# Spatial and temporal changes in choroid morphology associated with long-duration spaceflight

**DOI:** 10.1101/2024.10.01.24314650

**Authors:** Charles Bélanger Nzakimuena, Marissé Masís Solano, Rémy Marcotte-Collard, Mark Richard Lesk, Santiago Costantino

## Abstract

**Purpose:** Amid efforts to understand spaceflight associated neuro-ocular syndrome (SANS), uncovering the role of the choroid in its etiology is challenged by the accuracy of image segmentation. The present study extends deep learning-based choroid quantification from optical coherence tomography (OCT) to the characterization of pulsatile and topological changes in the macular plane and investigates changes in response to prolonged microgravity exposure.

**Methods:** We analyzed OCT macular videos and volumes acquired from astronauts before, during and after long-duration spaceflight. Deep learning models were fine-tuned for choroid segmentation and combined with further image processing towards vascularity quantification. Statistical analysis was performed to determine changes in time-dependent and spatially averaged variables from preflight baseline.

**Results:** For 12 astronauts with a mean age of 47 *±*9 years, there were significant increases in choroid thickness and luminal area (LA) averaged over OCT macular video segments. There was also a significant increase in pulsatile LA. For a subgroup of 6 astronauts for which inflight imaging was available, choroid volume, luminal volume and choroid vascularity index over the macular region all increased significantly during spaceflight.

**Conclusions:** The findings suggest that localized choroid pulsatile changes occur following prolonged microgravity exposure. They show that the choroid vessels expand in a manner similar to the choroid layer across the macular region during spaceflight, with a relative increase in the space they occupy. The methods developed provide new tools and avenues for studying and establishing effective countermeasures to risks associated with long-duration spaceflight.

## Introduction

Following prolonged exposure to microgravity, astronauts present with distinct neuro-ophthalmic findings collectively referred to as spaceflight associated neuroocular syndrome (SANS)^27^; ^35^. The signs defining SANS comprise unilateral and bilateral optic disc edema, posterior globe flattening, choroidal and retinal folds, hyperopic refractive error shifts, retinal nerve fiber layer infarcts, increased peripapillary total retinal thickness from the Bruch’s membrane opening to 250 µm and increased cerebrospinal fluid volume in optic nerve sheaths^16; 17; 38; 18; 21; 27; 35; 41^. While there have not been reports of permanent vision loss as a result of SANS, severe optic disc edema could damage visual pathways to the brain, and it has been identified as the chief risk associated with SANS^38^. Choroidal tissue undergoes acute variations in space over time^32^, and finite element modelling of the posterior segment of the eye suggests that increased pulsatile choroid volume fluctuation may elevate strains in prelaminar neural tissue. The increased strains could in turn lead to the development of edema^5; 35^.

Pulsatile choroid volume fluctuation has been used to compute ocular rigidity and evaluate its response to long-duration spaceflight^45^. The effect of microgravity exposure on pulsatile choroid volume fluctuation itself has not yet been scrutinized. However, there is evidence that the choroid expands as soon as it is exposed to microgravity^44; 21; 35^, and the peripapillary choroid remains thicker than the pre-spaceflight baseline for at least 30 days and up to 90 days after landing^21; 35^. The view that choroidal swelling may be in part responsible for the hyperopic shifts and choroidal folds observed in SANS^24; 16; 17; 18^, and the possible link between altered choroid pulsatility and edema warrant seeking a refined characterization of choroidal changes in response to microgravity.

The development of optical coherence tomography (OCT) with enhanced depth imaging has made possible the visualization of details of the choroid in most imaged subjects^32^ and enabled the performance of reproducible biometric measurements^52; 32; 14^. Several means of quantifying the morphology of the choroid from OCT representations have been devised^42^. Albeit routinely resorted to in clinical research contexts, manual segmentation of the choroid is labor-intensive^46; 55; 56^ and ill-suited to the considerable volume of data produced by OCT devices, entailing a pace vastly exceeded by recent automated approaches^55^. The automated choroid semantic segmentation landscape has evolved from the use of various classical image processing methods^3^; ^28^ to the application of deep neural networks^56^. Under the supervised deep learning paradigm, the transformer architecture has demonstrated superior capabilities compared to convolutional neural networks when applied to computer vision tasks including segmentation^37^; ^47^.

Within the context of choroid segmentation, a few methods enable the differentiation of vasculature lumen and walls. It is now possible to identify, within a single OCT B-scan, the luminal area (LA) and to calculate choroidal vascularity index (CVI), defined as the fraction of LA over the total choroid area. However, the successful application of such parameters is highly dependent upon the accurate delineation of the choroid^1^.

There is a correlation between pulsatile choroid volume fluctuation and ocular rigidity^39^, and ocular rigidity has been reported to decrease in response to microgravity exposure^44^; ^45^. We hypothesized that structural choroid vascular changes in the macular region could be quantified: we analyzed OCT-based spatial and temporal changes in the choroid morphology of astronauts during and following long-duration space missions.

For the present work, we trained deep learning models to reliably produce choroid semantic segmentations in macular images from OCT videos and OCT volumes acquired from astronauts. We complemented our segmentation techniques with vascularity quantification, enhancing the ability to characterize the state of the choroid and its variations. We extended the capability of our approaches to OCT image timeseries and OCT volume reconstructions. For video acquisitions, we compared pre- and post-spaceflight quantifications and revealed significant changes in time-dependant variables. For volumes, we captured substantial mapping changes over several timepoints in a cohort of astronauts exposed to microgravity.

## Methods

### Datasets

The study was approved by the institutional review boards of the Maisonneuve-Rosemont hospital and NASA, and the clinical research ethics committee of Université de Montréal, and it was conducted according to the Declaration of Helsinki and its amendment. Two datasets were used (see table S1 for a summary). All acquisitions were performed using spectral-domain OCT (Spectralis OCT2; Heidelberg Engineering, Heidelberg, Germany). Study participation required not having received any previous glaucoma-related surgeries and none were reported. No ocular pathologies were identified prior to spaceflight in any of the subjects participating in the study based on OCT and fundus examination.

OCT macular videos were obtained through a specialized feature complementary to the OCT device’s acquisition software which was provided to us by Heidelberg Engineering. The videos were acquired pre- and post-spaceflight from both eyes in the Enhanced Depth Imaging mode, with up to 5 videos per eye at each timepoint. Each frame has dimensions 768 × 496 pixels, approximately corresponding to 8.4 × 1.9 mm, and each video is roughly 1 minute in length. Each OCT video was acquired along a macular plane segment centered at the fovea. The segment was rotated bidirectionally to within 15-45 degrees from the horizontal (nasal to temporal direction) about the fovea, based on a trained operator’s visual assessment of the quality of the displayed frame. The operator’s quality optimization was performed at each acquisition independently. Video acquisitions had a specified automatic real time frame averaging of 5 or 10 frames. A resting heart rate measurement was also obtained concurrently with video acquisition.

For the analysis of changes in 3-dimensional topology due to microgravity exposure, we used a set of OCT macular volumes composed of 97 or 193 B-scans. The macula block setting was used with placement over the fovea via the anatomic positioning system for each acquisition. For each timepoint, up to 2 volumes per eye were obtained in the Enhanced Depth Imaging mode. All frames in the set of OCT macular volumes measure 512 × 496 pixels, which physically approximates to 5.6 × 1.9 mm.

### Segmentation training and performance evaluation

SegFormer models^54^ pretrained on the ImageNet database were retrained directly on both datasets towards choroid segmentation. For OCT videos, training data was gathered by randomly selecting 10 B-scans per movie for a total training dataset of 1070 B-scans where the choroid region was manually labelled by one trained grader and binarized. For OCT volumes, we devised a strategy to ensure adequate representation of the distribution of B-scans corresponding to each subject’s eye in the training data. The image set was split into 5 groups along the slow acquisition direction, and 4 B-scans from each group were randomly selected for a total of 20 per volume. For each volume, 4 B-scans out of 20 were manually labelled by one trained grader and binarized. The locations of the labelled B-scans were subsequently shifted as shown in figure S9 resulting in a training dataset of 260 B-scans.

The training of the SegFormer models was achieved through the IoU loss. For binary segmentation, which was used exclusively in the present study, the IoU loss equation is given by

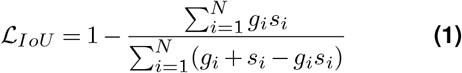

where *ℒ* _IoU_ is the IoU loss, N is the number of pixels in an image, s_i_ is the predicted value of the i^th^ pixel in an image, and g_i_ is the targeted ground truth value of the i^th^ pixel in an image.

The SegFormer models were trained using the Adam optimizer. A 0.0001 learning rate was used as well as early stopping based on validation loss for both Seg-Former models. No data augmentation strategy was carried as part of training the SegFormer models. All training was implemented on a Windows 11 computer equipped with a NVIDIA GeForce RTX 2060 Graphics Processing Unit (GPU). Training of the SegFormer transformer models was completed on Python (v3.10.0) using PyTorch Lightning (1.8.6).

Post-processing was implemented to ensure continuous layer boundaries and minimize the effect of isolated false positive regions (details are provided in Supplementary Information C).

We implemented 5-folds cross-validation on the videos and volumes datasets independently. The training data was arranged 5 different ways, in each instance splitting the whole into training, validation and test sets. For each fold, 10 % of B-scans were assigned to the validation set, and 10 % to the test set. The training and validation sets were used for model training and the test sets were used for performance evaluation.

The test set 5-fold cross-validation performance for both the videos and volumes datasets was evaluated based on similarity metrics, as well as choroid boundaries mean absolute error (MAE) and choroid thickness difference (TD) between the manually segmented and corresponding automatically segmented frames. The equations for MAE and TD are provided below

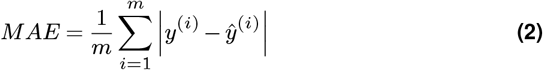

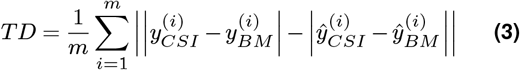

where m is the B-scan width and y^(i)^ and ŷ^(i)^ are the inferred and manually labeled vertical boundaries of Bruch’s membrane (BM) or the choroid-sclera interface (CSI). Sørensen–Dice coefficient complement (Dice^C^) and Jaccard coefficient complement (Jaccard^C^) were used as similarity metrics. For our study, Dice^C^ and Jaccard^C^ of 0 indicate perfect overlap between the manually and automatically segmented regions, and 1 no overlap. We also defined mathematical expressions to identify inaccurate segmentation and segmentation failure associated with subtle and gross segmented boundary aberrations, respectively (equations are provided in Supplementary Information D).

Beyond choroid segmentation, we produced masks of LA to calculate CVI (details are provided in Supplementary Information E). CVI was calculated as LA divided by total choroid layer area. The LA processing performance was assessed on the test set of selected Seg-Former models for both videos and volumes using similar methods as for choroid segmentation performance evaluation. Additional test set performance evaluations relating to choroid layer and lumen segmentation are provided in Supplementary Information I. Subsequent temporal and spatial analyses were performed on the complete datasets including the training data.

### Temporal analysis

Inference was only performed on B-scans which met noise specifications based on OCT device manufacturer defined metrics. The Spectralis OCT provides a quality score in dB based on a signal-to-noise ratio estimate^2^. Only B-scans with a quality score equal to or greater than 24 dB (0 dB-40 dB score range; 0 dB indicating no signal and 40 dB indicating excellent quality) and with an automatic real time frame averaging number greater or equal to 2 were considered.

We also implemented our own signal-to-noise ratio and excluded B-scans with a ratio beneath 0.55. The signal-to-noise ratio equation we used is provided below

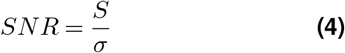

where SNR is the signal-to-noise ratio, S is the mean and *σ* is the standard deviation of the intensities in a B-scan. B-scans for which the inferred choroid region was located within 10 % of the superior or inferior vertical edge of the image were excluded. Single B-scans with an inferred choroid area (CA) departing from the average of the manually segmented frames by greater or equal to 20 % were also excluded.

OCT videos were unevenly sampled and frequency analysis was performed with Lomb–Scargle periodograms and spectrograms generated over a 0.1-4 Hz frequency range (transforms implementation details and calibration test results are provided in Supplementary Information G). Movies with less than 50 good quality frames were discarded. To prevent large gaps in a video, we preserved only the longest video segment for which no zone of excluded frames amounting to greater than 1 s was present. Pulsatile CT, LA and CVI (ΔCT, ΔLA, ΔCVI) were computed as done by Beaton et al^3^. First, the locations of peaks and valleys across time series were obtained, where the minimum distance between them is given by the expression

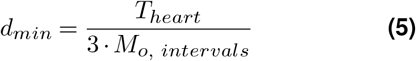

where d_min_ is the minimum distance between peaks and between valleys, T_heart_ is the heart period and M_o, intervals_ is the mode of the 1D array of intervals between each element in a time series. Having obtained peaks and valleys locations, consecutive peaks between valleys and consecutive valleys between peaks were discarded. ΔCT, ΔLA and ΔCVI were then defined as the difference between the median of all remaining peaks and the median of all remaining valleys. Each quantification result was divided into macular ETDRS subfields (center; nasal inner; temporal inner; nasal outer; temporal outer), in addition to a global level value.

A video was excluded entirely if more than 150 B-scans were under the signal-to-noise specification or met the edge exclusion criterion. Furthermore, a video with ΔCT far exceeding previously reported upper physiological values in any region was not kept. The scheme for exclusion based on ΔCT was implemented through the following equation

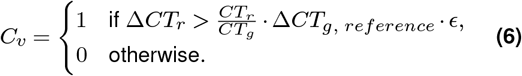

where C_v_ is the choroid pulsatility-based video exclusion criterion (with 1 indicating exclusion and 0 indicating inclusion), ΔCT_r_ and CT_r_ are global or ETDRS region measurements, CT_g_ is the global measure, ΔCT_g, reference_ is a global constant of 20 µm and *ϵ* represents an excess threshold of 3. Having obtained temporal choroid quantifications, we determine statistical significance of differences between pre- and post-spaceflight timepoints.

### Spatial analysis

For OCT volumes, the automatic real time averaging setting was 15 frames per B-scan. Following initial segmentation, smoothing of the surface of the CSI across each macular volume was implemented using a graph search algorithm similar to the one described by Mazzaferri et al^28^. The graph search algorithm was applied to each one of the volume’s cross-sectional image along the fast acquisition direction.

After delineating the region between the BM and a smoothed CSI, choroid volume (CV), luminal volume (LV) and CVI were quantified (CT and LT are included in Supplementary Information N). In addition to a global level value, the result of each quantification was split into macular ETDRS subfields (center; superior inner; nasal inner; inferior inner; temporal inner; superior outer; nasal outer; inferior outer; temporal outer).

For better visualization, we created square maps of CT, LT and CVI of 512 × 512 pixels by interpolating the total number of B-scans of every OCT volume and we analysed variation across timepoints. We determined the statistical significance of spatial choroid measurement differences between timepoints.

### Repeatability

We evaluated the repeatability of choroid measurements using sets of OCT videos and volumes obtained in the same session by gathering the significance of differences. We calculated intraclass correlation coefficient and standard error of measurement between these measures. Our repeatability evaluations are described, and complete results are provided in Supplementary Information J and K.

### Statistical analyses

Linear mixed models were used to determine the statistical significance of differences between measurements. For each linear mixed model, timepoints (or scans for repeatability analyses) were designated as the fixed effect. Subjects and eyes nested under each subject were specified as random effects^4^. The linear mixed models were implemented in R (v4.3.1) using the lme4 package (1.1.33). Significance values were gathered using the Kenward-Roger approximation^20^.

## Results

### SegFormer-based choroid segmentation

To show choroid morphology changes in response to microgravity exposure, we retrained SegFormer models directly on astronaut OCT macular videos and volumes. We combined our segmentation with a simple strategy to analyze vascular changes. We manually labeled a 2 % subset of the available images and assessed our pipeline’s test set performance using different metrics.

We leveraged a dataset of OCT macular videos to compare pre- and post-spaceflight time-dependant variables obtained from astronauts. We used images centered at the macula from 12 individuals, 9 male and 3 female of average age 47 *±*9 yo (1 subject of an initial 13 was excluded based on video filtering criteria). There were 8 astronauts who did not have prior space-flight experience, 3 who had completed Space Shuttle short-duration missions, and 3 who had previous long-duration spaceflight experience (*>* 4 months^19^). All pre-flight videos were obtained within a 9 to 1 month before spaceflight window and all postflight videos within 2 to 30 days after spaceflight.

Figure 1a and d display illustrations of 3 individuals’ manual and automated segmentations showing moderate-high to high correspondence for choroid and vascular lumen obtained from OCT videos. Mean Dice^C^ and mean Jaccard^C^ for the OCT videos test set (figure 1b) were 0.044 *±*0.02 and 0.083 *±*0.03, respectively. Figure 1c provides segmentation mean absolute error (MAE), expressed as the deviation of the boundary (in pixel units) from the labeled traces. It also shows thickness difference (TD) which represents the deviation of the layer thickness (in pixels) from the labeled layer. Test set BM and CSI MAE were 0.93 *±*0.3 pixels and 6.2 *±*2.7 pixels, respectively, and TD was 6.1 ± 2.7 pixels. Test set LA mean Dice^C^ and mean Jaccard^C^ were 0.32 *±*0.1 and 0.48 *±*0.09, respectively (figure 1e).

**Figure 1.**
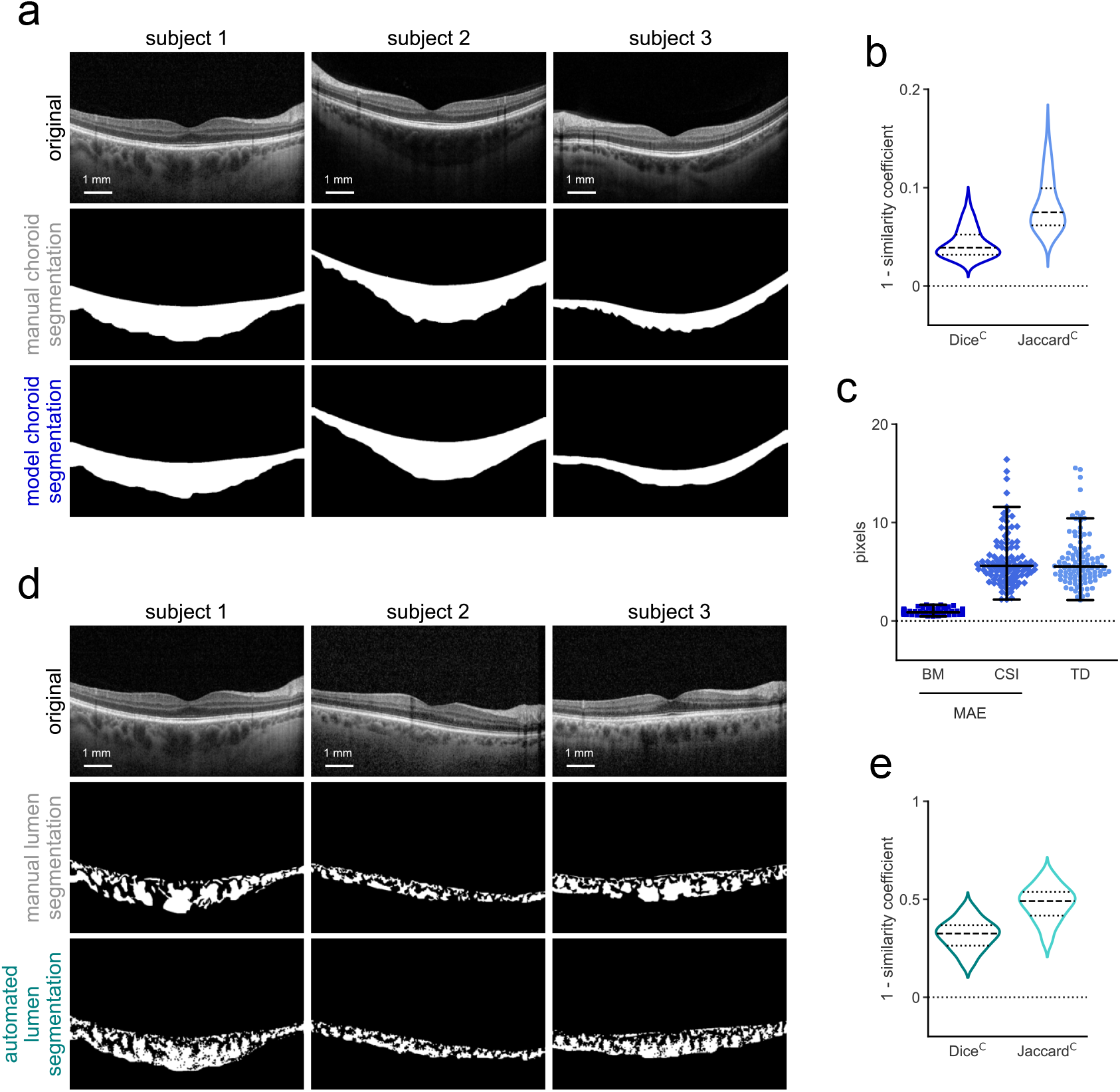
Choroid layer and lumen segmentation performance evaluation for OCT videos. (**a**) Original OCT video B-scans and binary choroid layer segmentation masks for 3 subjects obtained manually and through model inference. (**b**) Violin plots displaying test set Dice^C^ and Jaccard^C^ between manual and model choroid layer segmentations. (**c**) Swarm plots of test set BM and CSI mean absolute errors and average choroid thickness difference between manual and model choroid layer segmentations. (**d**) Original OCT video B-scans and binary lumen segmentation masks for 3 subjects obtained manually and automatically. (**e**) Violin plots displaying test set Dice^C^ and Jaccard^C^ between manual and automated lumen segmentations.

The performance was similar for the B-Scans from OCT volumes (manual and automated segmentations are shown in figure 2a and d). This second set contains OCT volumes centered at the macula from 6 of the 12 astronauts featured in on-Earth videos, 5 males and 1 female, which were of average age 48 *±*9 yo. There were 4 astronauts who did not have prior space-flight experience, 1 who had completed Space Shuttle short-duration missions, and 2 who had previous long-duration spaceflight experience. The volumes were obtained at up to 6 different timepoints before launch (launch - 21 to 18 months, launch - 9 to 6 months), during flight and before return (launch + 30 days, launch + 90 days, return - 30 days) and following spaceflight (return + 1 to 3 days). Test set choroid segmentation mean Dice^C^ and mean Jaccard^C^ for OCT macular volumes were 0.057 *±*0.02 and 0.11 *±*0.04, respectively (figure 2b). Test set BM and CSI MAE for the same fold were 1.2 *±*0.3 pixels and 8 *±*4.2 pixels, respectively, and TD was 7.9 ± 3.9 pixels (figure 2c). Test set LA mean Dice^C^ and mean Jaccard^C^ were 0.39 *±*0.07 and 0.55 *±*0.07, respectively (figure 2e). Additional choroid layer segmentation and lumen segmentation performance evaluations are provided in supplementary figures S10 and S11.

**Figure 2.**
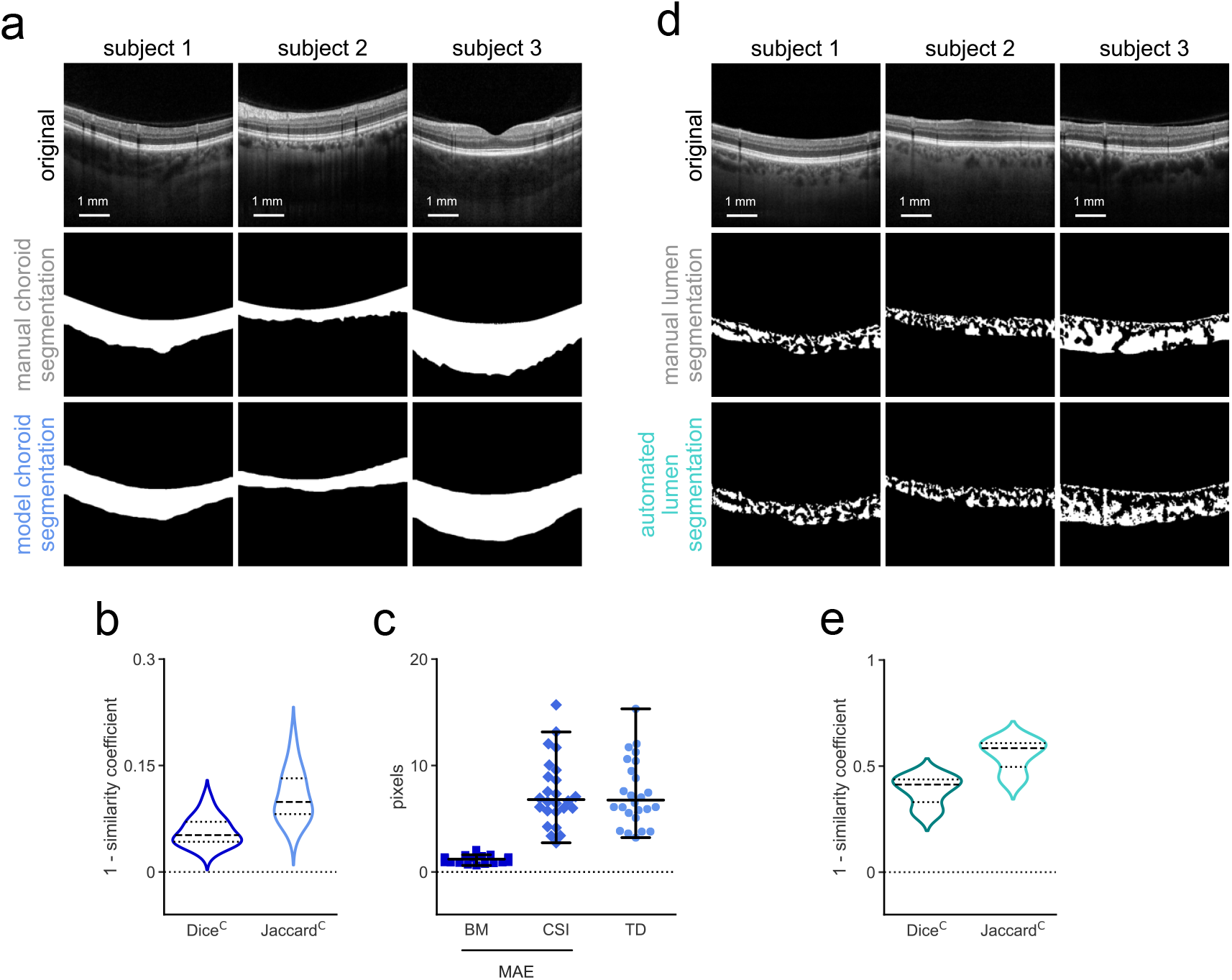
Choroid layer and lumen segmentation performance evaluation for OCT volumes. (**a**) Original OCT volume B-scans and binary choroid layer segmentation masks for 3 subjects obtained manually and through model inference. (**b**) Violin plots displaying test set Dice^C^ and Jaccard^C^ between manual and model choroid layer segmentations. (**c**) Swarm plots of test set BM and CSI mean absolute errors and average choroid thickness difference between manual and model choroid layer segmentations. (**d**) Original OCT volume B-scans and binary lumen segmentation masks for 3 subjects obtained manually and automatically. (**e**) Violin plots displaying test set Dice^C^ and Jaccard^C^ between manual and automated lumen segmentations.

### Quantification of choroid pulsatile changes

We investigated the presence of spaceflight-related changes in choroid pulsatility. For this, as we have done in the past^3^, we obtained videos of OCT images of about 1 minute, acquired at 14 Hz. The builtin eye tracker of the OCT device was used to assure that the same location in the macula was sampled throughout the video. The acquisition was automatically halted when a patient moved, only resuming after the laser was repositioned. Our choroid quantification approaches were implemented on the videos, yielding timeseries which displayed morphological changes at the frequency of the heart. Because images are not sampled at a constant rate, Lomb–Scargle periodograms and spectrograms of normalized power were expected to show peaks in the heart rate vicinity. Detection of those peaks, as illustrated in figure 3d-f, serves as quality control for segmentation. Narrow and prominent maxima can often be seen for CT, LA and CVI (also figures S18 and S19, second and third columns).

**Figure 3.**
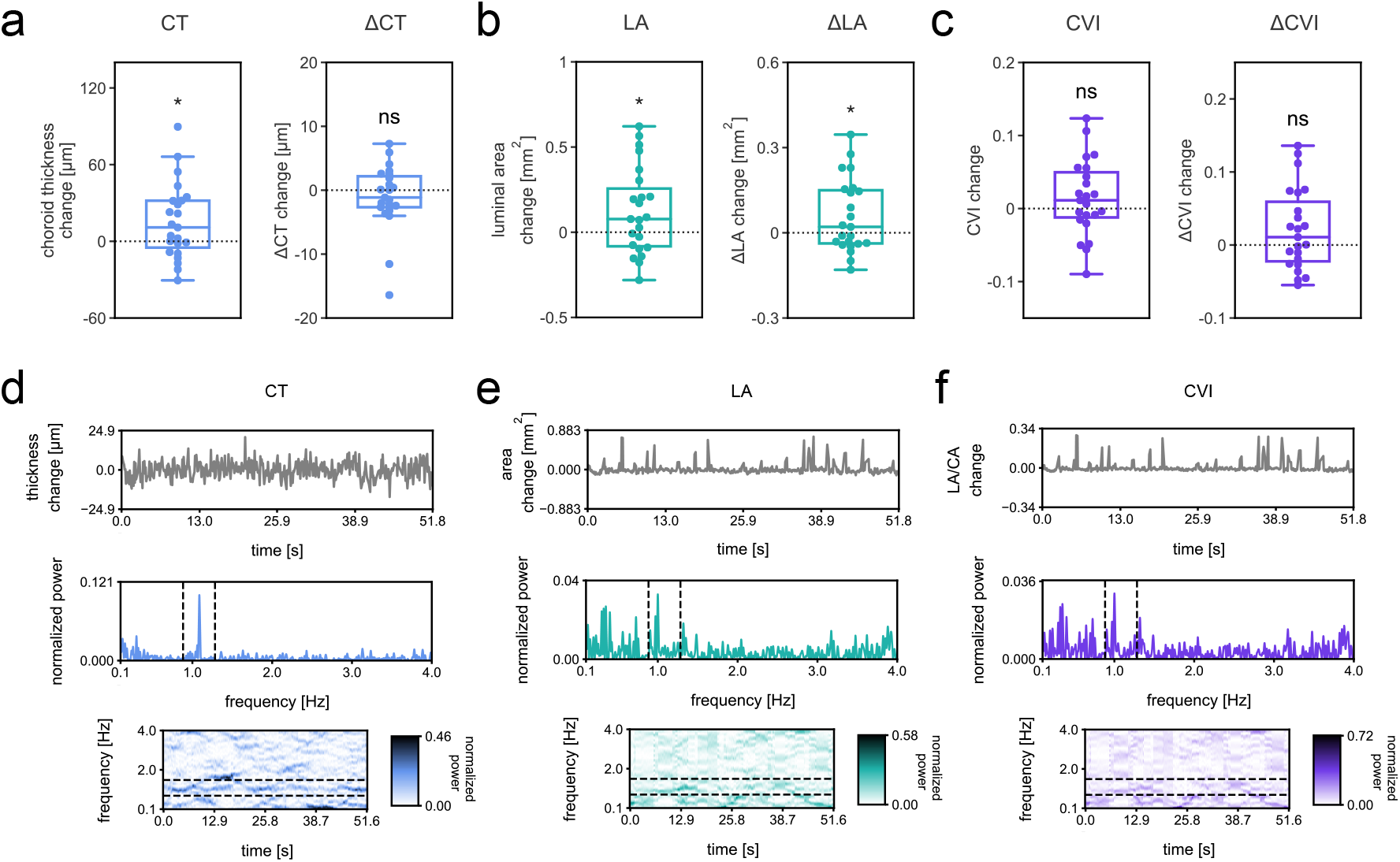
Temporal choroid pulsatile changes. Difference box plots between post- and preflight for: (**a**) CT and ΔCT measurements obtained over the selected segments for all astronaut eyes, (**b**) LA and ΔLA, and (**c**) CVI and ΔCVI. ns nonsignificant, *P *<* 0.05, linear mixed model. Selected segment timeseries, frequency and joint time-frequency transforms for one subject’s eye acquisition displaying the behavior of (**d**) CT, (**e**) LA, and (**f**) CVI. Top row features timeseries, second row features the Lomb–Scargle periodograms and third row features the Lomb–Scargle spectrograms. Dashed lines on the periodograms and spectrograms show values *±* 20 % and *±* 40 % of the oximeter measured heart rate value, respectively.

We found a global CT averaged over movie durations significant increase of 15 *±*30 µm or 5.3 % (linear mixed model, P = 0.02) from pre-to postflight, and global LA significant increase of 0.12 *±*0.3 mm^2^ or 7.9 % (linear mixed model, P = 0.03) (figure 3a and b), but no change in global CVI (figure 3c). For the analysis of the amplitude of pulsatile fluctuations, no significant differences in ΔCT or ΔCVI were found globally (figure 3a and c). As shown in figure 3b, global ΔLA did show a significant increase from pre-to postflight of 0.058 ± 0.13 mm^2^ or 25 % (linear mixed model, P = 0.03). We provide baseline pre-spaceflight results of our choroid quantification methods for OCT videos from astronaut eyes, including their ETDRS subfield equivalents in supplementary table S6. The ETDRS subfield equivalents of the measures displayed in figure 3a-c are provided in supplementary figure S17, and significance in table S8.

### Spatial choroid changes associated with long-duration spaceflight

We extended our choroid quantification techniques to OCT volumes, for which large variations were expected in astronauts. Each volume is made up of 97 or 193 B-scans. In addition to obtaining cross-sectional image segmentation performance, we generated maps exposing choroid thickness topography and vascular patterns. Choroid vessel patterns were readily observable on LT and CVI maps in OCT volumes (figure 4d). For all eyes, there was a correspondence between CT and LT maps, with thicker choroid regions generally translating to thicker luminal regions. Different degrees of retinal vessel shadow artifacts were discernable in most scans.

**Figure 4.**
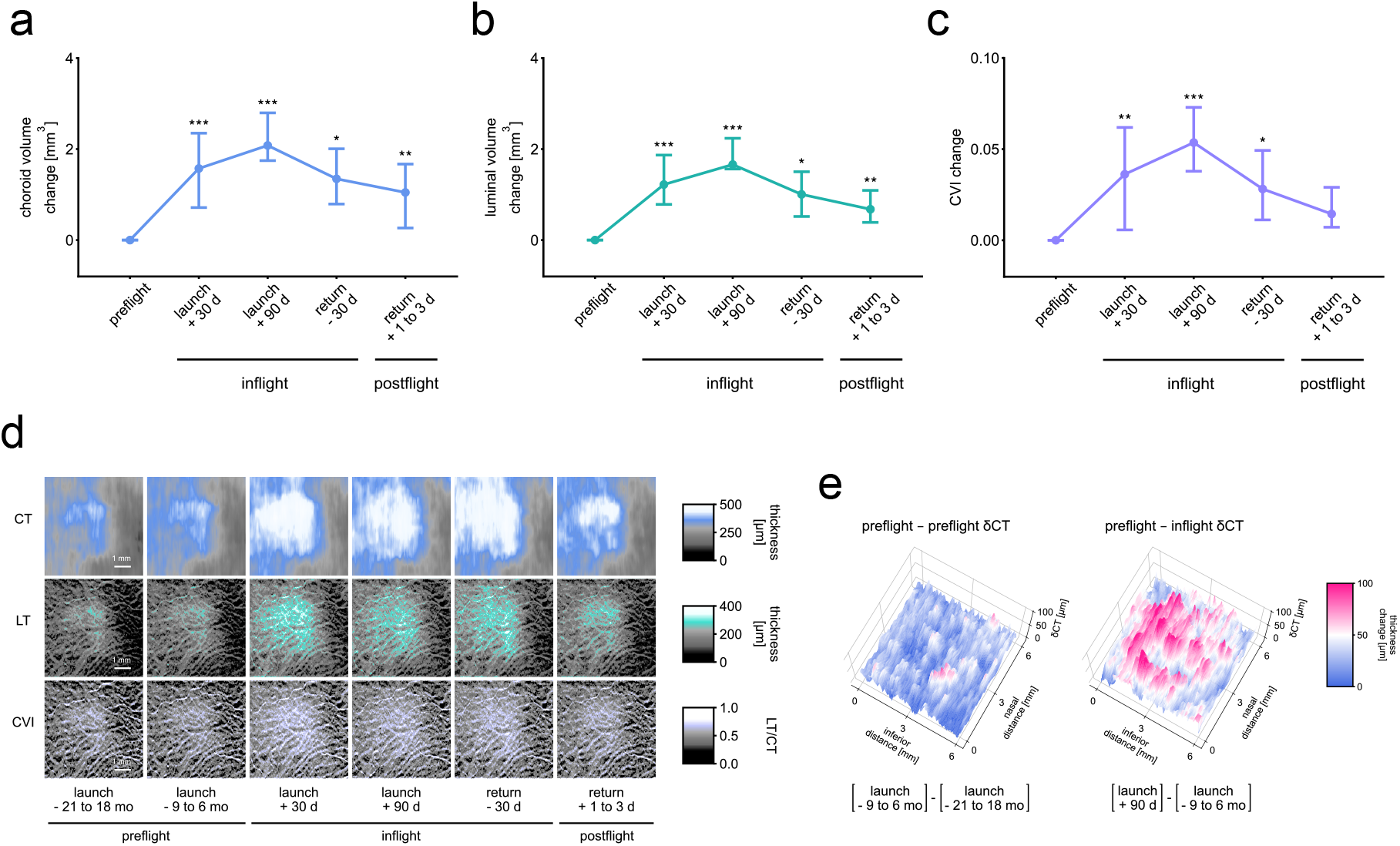
Spatial choroid changes. Line charts showing changes over preflight, inflight and postflight timepoints for all astronaut eyes for: (**a**) choroid volume, (**b**) luminal volume, and (**c**) CVI. Error bars show the difference interquartile range and dots show the mean difference. (d days). *P *<* 0.05, **P *<* 0.01, ***P *<* 0.001, linear mixed model. (**d**) Maps of CT, LT and CVI over preflight, inflight and postflight timepoints for one subject’s eye. The top row displays CT maps, the middle row displays LT maps, and the bottom row displays CVI maps. (d days, mo months). (**e**) Surface plots of the difference between two preflight CT maps (launch - 21 to 18 months and launch - 9 to 6 months) (left), and of the difference between preflight and inflight CT maps (launch - 9 to 6 months and launch + 90 days) (right) for the same subject’s eye acquisitions. (*δ*CT choroid thickness change).

When comparing individual timepoints with preflight, there were significant differences for global CV and LV (figure 4a and b) for all measurements. The same comparison for global CVI measurements indicated significant differences for inflight timepoints and normal values for return + 1 to 3 days (figure 4c). We provide baseline results of our choroid quantification methods for OCT volumes from astronauts, including their ET-DRS subfield equivalents in supplementary table S7. The ETDRS subfield equivalents of measures displayed in figure 4a-c are provided in Supplementary Information N, and significance values in supplementary tables S9-S12.

## Discussion

In addition to an increase in CT within the macular region during the month that follows long-duration space- flight, we found an increase in the area corresponding to the lumen. Our results also indicate that while pulsatile fluctuation in CT did not change compared to preflight when measured within 30 days of return, the amplitude of pulsatile fluctuation in LA increases. During missions, we observed a global increase in the space occupied by lumen relative to the whole choroid layer across the macular plane.

Consistent with our results, a study which measured macular CT preflight and during spaceflight showed a 35 µm CT increase^7^. Previous reports demonstrated similar increases in peripapillary CT. Laurie et al found a mean peripapillary CT increase of 27 µm during space- flight compared to preflight^12^. Macias et al observed progressive peripapillary choroid thickening over the course of long-duration spaceflight, with a mean increase of 43 µm at 150 days^21^. In line with our findings for CT at the macula, they also showed still significantly increased peripapillary CT at 30 days following return to Earth. Unlike the present work, previous studies were limited to measuring CT from a single image. Here we complement previous reports by adding maps of thickness change, as well as an analysis of how lumen and CVI evolve and their response to cardiac pulsation.

Several mechanisms have been proposed to account for increased CT upon microgravity exposure. They include cephalad fluid shift resulting in venous congestion and accumulation of blood in the choroid^22; 16; 18^ as well as persistent buildup of choroidal interstitial fluid in the peripapillary area^51; 35^. In line with the posited cephalad fluid shift mechanism, a mathematical model of the cardiovascular response to long-duration spaceflight predicted increased upper body blood volume^6^.

Determining the source of optic disc edema observed in SANS has been described as challenging using available technology and approaches to data analysis^21^. The results of finite element modelling suggest a link between choroid anatomy, increased pulsatile choroid volume fluctuation, prelaminar neural tissue strains and the development of edema^5; 35^. Our study directly shows increased amplitude of pulsatile fluctuation of the space occupied by choroid vessels at the macula after space- flight. The finding opens the possibility of investigating whether a ΔLA elevation can also be observed in the peripapillary region. A relationship could in turn be explored between the latter and peripapillary retinal thickness increase in SANS. Pulsatile deformation of the optic nerve head may be assessed using OCT^43^ and could also be studied in relation to peripapillary choroid pulsatile changes, helping substantiate finite element modelling predictions.

Studies have linked the development of edema in SANS with single nucleotide polymorphisms (SNPs)^57^; ^58^. A lower enzyme functional activity in the 1-carbon pathway can increase B-vitamins requirements and the risk of vitamin insufficiency^59^. The insufficiency may lead to the generation of reactive oxygen species^9^; ^30^, with a deleterious effect on interendothelial junctions and a rise in protein and fluid leakage into the interstitial space^8^; ^31^. A significant association has been found between choroid thickness change in SANS and peripapillary retinal thickness increase^41^, an imaging biomarker of disc edema^29^; ^33^, and common factors may underly variation in their manifestation. Specifically, both individual differences in posterior segment anatomy^5^ and SANS-relevant SNPs^57^; ^58^ could contribute to the degree of choroid morphological changes observed.

The application of lower body negative pressure has been explored as a means of mitigating the effects of cephalad fluid shift in SANS. While lower body negative pressure was shown to decrease intraocular pressure in astronauts during spaceflight, no change in macular CT was observed^7^. Numerical modelling suggests that under microgravity conditions, less rigid vessels are subjected to increased transmural pressure and greater than normal volume^10^. An artificial gravity experiment achieved partial alleviation of mice retinal tissue damage associated with weightlessness exposure^25; 48^. The effects of artificial gravity on the state of the choroid have not been explored, and the need for precise and reliable methods of assessing SANS-related eye alterations to validate its effectiveness as a countermeasure has been emphasized^48^. The approaches we developed for the measurement of pulsatile and volume choroid changes represent a step in this direction.

Head-down tilt bed rest has been explored as a SANS analog and towards studying the effect of countermeasures. There have been conflicted reports about the impact of head-down tilt bed rest on CT in the peripapillary and macular regions. Some studies have shown significantly increased CT in response head-down tilt bed rest^40; 11; 15^, with others showing no CT alteration^36; 13^. Choroid changes observed were generally lower in the analog than those resulting from microgravity exposure^13; 34^, which is consistent with our observation of the differences reached at 90 days following launch. Unlike during spaceflight, significantly increased intraocular pressure has been reported in head-down tilt bed rest studies^40; 11; 36; 13^. When comparing head-down tilt bed rest and prolonged microgravity exposure with respect to the development of optic disc edema, differences in the combined effects of posterior segment anatomy, pulsatile choroid volume fluctuation and IOP may be at play, as predicted by finite element modeling^5^.

A relationship between venous congestion in SANS and raised intracranial pressure has been proposed^23^. In contrast with the pervasiveness of choroid enlargement in space, the manifestation of visible choroidal folds may depend on intracranial hypertension and individual microanatomical variation. From a biomechanical perspective, an early exploration of ocular rigidity in a private astronaut suggested that it decreases following spaceflight^44^, and this was recently confirmed in professional astronauts^45^. It has been suggested that thickening of the choroid could extend scleral collagen and modify its properties resulting in the observed lowered postflight ocular rigidity.

Pulsatile choroid volume fluctuation correlates inversely with the measurement of ocular rigidity performed using an invasive procedure^39^. Combined with additional measurements, our choroid layer segmentation method has the potential to improve upon a key element in the computation of ocular rigidity and delve deeper into its relationship to SANS. Furthermore, the significant post-flight global and localized increases in ΔLA we observed could be associated with increased choroidal vessel compliance following sustained greater than normal volume during long-duration spaceflight. The techniques we described here offer novel approaches as well as opportunities to explore SANS etiology, and to implement effective strategies against risks linked with extended space travel.

Our study is not without some limitations, and they should be highlighted. First, our results suggest that there is considerable variability in model predictions, and they systematically underestimate the size of the choroid space. In spite of the limitation, the BM and CSI MAEs, choroid TD and Dice^C^ for OCT videos and volumes were comparable with reported values obtained using similar methods on myopic children and adults ^55^; 49. Similar performance was also achieved when comparing with the segmentation of healthy choroids using convolutional neural networks ^26^; ^53^. Manual and model segmentations produced well correlated measurements across the available range of choroid thicknesses. Despite the correlations, the CSI can be challenging to identify visually in very thick choroids, and this could have adversely impacted our results. Unlike with learned choroid segmentation, with our automated lumen quantification approach there was a systematic area overestimation. Perhaps, this is owing at least in part to the difficulty of capturing very small luminal detail by hand. Using a scheme previously described ^50^, luminal area segmentation achieved moderate-high similarity. Nevertheless, it did not perform as well as choroid segmentation and showed sizable variability.

In addition to segmentation performance considerations, our sample size is modest, and a larger number of participants would have enabled us to achieve a superior level of power. The significant changes we observed should be interpreted cautiously, and with consideration of our repeatability assessment. The time interval between preflight acquisitions and launch was not considered as a fixed effect in the linear mixed models when analyzing OCT videos. We recognize that the exclusion of this variable as a fixed effect represents a limitation of our approach. Owing in part to our sample size constraint, participants were not grouped based on their genetic profile or flight experience. We could address these limitation in the future by using the approaches we developed to determine whether individuals with SANS-relevant SNPs or prior flight experience demonstrate the same or greater choroid morphology changes in response to microgravity. Importantly, their relationship with peripapillary retinal thickness alterations could be elucidated. A control group of astronauts which would remain on Earth was not included as part of our study. In support of our approach, no significant differences were found in our evaluation of repeatability. There was also generally a marked distinction in the magnitude of the difference between two preflight timepoints, and when comparing our baseline with subsequent timepoints.

Lastly, there were additional factors which could have affected our results. The time of day of initial and subsequent OCT acquisitions was not always consistent across timepoints and this could have impacted our findings. In most instances where significance was found, changes in response to microgravity exposure reached magnitudes beyond what could reasonably be anticipated from diurnal fluctuations alone. The results corresponding to changes in metrics obtained from OCT videos and OCT volumes were also not adjusted for axial length. We acknowledge the possibility that establishing a relationship between choroid-based metrics and axial length for our data could help reveal important physiological insights.

## Conclusions

In conclusion, neuro-ophthalmic findings in spaceflight associated neuro-ocular syndrome (SANS) including optic disc edema constitute important risks for astronauts and their missions. Weightlessness results in rapid choroidal expansion which may partially account for concerning SANS findings. Macular region luminal area and the amplitude of its pulsatile fluctuation were both increased compared to preflight within 30 days after long-duration spaceflight. During long-duration spaceflight, there is a global increase in both the choroid layer and the relative space occupied by lumen inside the choroid across the macular plane.

## Data Availability

All data produced in the present study are available upon reasonable request to the authors.

## Acknowledgements

Funding was provided by the Canadian Space Agency, the Canadian Institutes of Health Research, and the Fonds de Recherche en Ophtalmologie de l’Université de Montréal (FROUM). MMS received a scholarship from the Fonds de Recherche du Québec Santé. SC holds the Wolfe Professorship in Translational Research. We thank NASA and ESA for coordination and infrastructure.

## Notes

### Competing Interest Statement

The authors have declared no competing interest.

### Author Declarations

The IRB of the Maisonneuve-Rosemont hospital gave ethical approval for this work. The National Aeronautics and Space Administration IRB gave ethical approval for this work. The clinical research ethics committee of Université de Montréal gave ethical approval for this work.

### Summary of Updates

Clarifications were added to the methods and discussion. Segmentation performance was split into two figures. Network training and assessment descriptions were combined. An analysis of repeatability was added and includes an evaluation of whether differences between measurements from repeated acquisitions were statistically significant. Intraclass correlation coefficient and standard error of measurement were obtained for the same measures. Based on the results of repeatability analysis, criteria for the exclusion of whole videos were added and a datapoint was removed which made significance more robust. The acquisition time of day of OCT videos and volumes was added. A minor error in the statistical testing approach for spatial choroid changes which had a small effect on significance was fixed. Choroid temporal changes were normalized, presenting a difference in each measurement from baseline to improve visualization. A discussion of the variability in choroid and lumen segmentation performance was added and comparisons with previous reports were offered when available. All study limitations were consolidated into a dedicated section in the discussion.

## References

1. Rupesh Agrawal, Jianbin Ding, Parveen Sen, Andres Rousselot, Amy Chan, Lisa Nivison-Smith, Xin Wei, Sarakshi Mahajan, Ramasamy Kim, Chitaranjan Mishra, et al. Exploring choroidal angioarchitecture in health and disease using choroidal vascularity index. Progress in retinal and eye research, 77:100829, 2020.

2. Madhusudhanan Balasubramanian, Christopher Bowd, Gianmarco Vizzeri, Robert N Weinreb, and Linda M Zangwill. Effect of image quality on tissue thickness measurements obtained with spectral domain-optical coherence tomography. Optics express, 17(5):4019–4036, 2009.

3. L Beaton, J Mazzaferri, F Lalonde, M Hidalgo-Aguirre, D Descovich, MR Lesk, and S Costantino. Non-invasive measurement of choroidal volume change and ocular rigidity through automated segmentation of high-speed oct imaging. Biomedical optics express, 6(5):1694–1706, 2015.

4. Qiao Fan, Yik-Ying Teo, and Seang-Mei Saw. Application of advanced statistics in ophthalmology. Investigative ophthalmology & visual science, 52(9):6059–6065, 2011.

5. Andrew J Feola, Emily S Nelson, Jerry Myers, C Ross Ethier, and Brian C Samuels. The impact of choroidal swelling on optic nerve head deformation. Investigative ophthalmology & visual science, 59(10):4172–4181, 2018.

6. Caterina Gallo, Luca Ridolfi, and Stefania Scarsoglio. Cardiovascular deconditioning during long-term spaceflight through multiscale modeling. npj Microgravity, 6(1):27, 2020.

7. Scott H Greenwald, Brandon R Macias, Stuart MC Lee, Karina Marshall-Goebel, Douglas J Ebert, John HK Liu, Robert J Ploutz-Snyder, Irina V Alferova, Scott A Dulchavsky, Alan R Hargens, et al. Intraocular pressure and choroidal thickness respond differently to lower body negative pressure during spaceflight. Journal of Applied Physiology, 131 (2):613–620, 2021.

8. Hadi AR Hadi, Cornelia S Carr, and Jassim Al Suwaidi. Endothelial dysfunction: cardiovascular risk factors, therapy, and outcome. Vascular health and risk management, 1(3):183–198, 2005.

9. Zvonimir S Katusic. Vascular endothelial dysfunction: does tetrahydrobiopterin play a role? American Journal of Physiology-Heart and Circulatory Physiology, 281(3):H981– H986, 2001.

10. Mimi Lan, Scott D Phillips, Veronique Archambault-Leger, Ariane B Chepko, Rongfei Lu, Allison P Anderson, Kseniya S Masterova, Abigail M Fellows, Ryan J Halter, and Jay C Buckey. Proposed mechanism for reduced jugular vein flow in microgravity. Physiological Reports, 9(8):e14782, 2021.

11. Steven S Laurie, Gianmarco Vizzeri, Giovanni Taibbi, Connor R Ferguson, Xiao Hu, Stuart MC Lee, Robert Ploutz-Snyder, Scott M Smith, Sara R Zwart, and Michael B Stenger. Effects of short-term mild hypercapnia during head-down tilt on intracranial pressure and ocular structures in healthy human subjects. Physiological reports, 5(11): e13302, 2017.

12. Steven S Laurie, Stuart MC Lee, Brandon R Macias, Nimesh Patel, Claudia Stern, Millennia Young, and Michael B Stenger. Optic disc edema and choroidal engorgement in astronauts during spaceflight and individuals exposed to bed rest. JAMA ophthalmology, 138(2):165–172, 2020.

13. Steven S Laurie, Scott H Greenwald, Karina Marshall-Goebel, Laura P Pardon, Akash Gupta, Stuart MC Lee, Claudia Stern, Haleh Sangi-Haghpeykar, Brandon R Macias, and Eric M Bershad. Optic disc edema and chorioretinal folds develop during strict 6° head-down tilt bed rest with or without artificial gravity. Physiological Reports, 9(15): e14977, 2021.

14. H Laviers and H Zambarakji. Enhanced depth imaging-oct of the choroid: a review of the current literature. Graefe’s Archive for Clinical and Experimental Ophthalmology, 252:1871–1883, 2014.

15. Justin S Lawley, Gautam Babu, Sylvan LJE Janssen, Lonnie G Petersen, Christopher M Hearon Jr, Katrin A Dias, Satyam Sarma, Michael A Williams, Louis A Whitworth, and Benjamin D Levine. Daily generation of a footward fluid shift attenuates ocular changes associated with head-down tilt bed rest. Journal of applied physiology, 129(5):1220– 1231, 2020.

16. Andrew G Lee, Thomas H Mader, C Robert Gibson, and William Tarver. Space flight– associated neuro-ocular syndrome. JAMA ophthalmology, 135(9):992–994, 2017.

17. Andrew G Lee, Thomas H Mader, C Robert Gibson, Tyson J Brunstetter, and William J Tarver. Space flight-associated neuro-ocular syndrome (sans). Eye, 32(7):1164–1167, 2018.

18. Andrew G Lee, Thomas H Mader, C Robert Gibson, William Tarver, Pejman Rabiei, Roy F Riascos, Laura A Galdamez, and Tyson Brunstetter. Spaceflight associated neuro-ocular syndrome (sans) and the neuro-ophthalmologic effects of microgravity: a review and an update. npj Microgravity, 6(1):7, 2020.

19. Stuart MC Lee, L Christine Ribeiro, David S Martin, Sara R Zwart, Alan H Feiveson, Steven S Laurie, Brandon R Macias, Brian E Crucian, Stephanie Krieger, Daniela Weber, et al. Arterial structure and function during and after long-duration spaceflight. Journal of Applied Physiology, 129(1):108–123, 2020.

20. Steven G Luke. Evaluating significance in linear mixed-effects models in r. Behavior research methods, 49:1494–1502, 2017.

21. Brandon R Macias, Nimesh B Patel, C Robert Gibson, Brian C Samuels, Steven S Laurie, Christian Otto, Connor R Ferguson, Stuart MC Lee, Robert Ploutz-Snyder, Larry A Kramer, et al. Association of long-duration spaceflight with anterior and posterior ocular structure changes in astronauts and their recovery. JAMA ophthalmology, 138(5): 553–559, 2020.

22. COL Thomas H Mader, C Robert Gibson, Michael Caputo, Norwood Hunter, Gerald Taylor, John Charles, and Richard T Meehan. Intraocular pressure and retinal vascular changes during transient exposure to microgravity. American journal of ophthalmology, 115(3):347–350, 1993.

23. Thomas H Mader, C Robert Gibson, Anastas F Pass, Larry A Kramer, Andrew G Lee, Jennifer Fogarty, William J Tarver, Joseph P Dervay, Douglas R Hamilton, Ashot Sargsyan, et al. Optic disc edema, globe flattening, choroidal folds, and hyperopic shifts observed in astronauts after long-duration space flight. Ophthalmology, 118(10): 2058–2069, 2011.

24. Thomas H Mader, C Robert Gibson, and Andrew G Lee. Choroidal folds in astronauts. Investigative Ophthalmology & Visual Science, 57(2):592–592, 2016.

25. Xiao W Mao, Stephanie Byrum, Nina C Nishiyama, Michael J Pecaut, Vijayalakshmi Sridharan, Marjan Boerma, Alan J Tackett, Dai Shiba, Masaki Shirakawa, Satoru Takahashi, et al. Impact of spaceflight and artificial gravity on the mouse retina: biochemical and proteomic analysis. International journal of molecular sciences, 19(9):2546, 2018.

26. Xiaoqian Mao, Yitian Zhao, Bang Chen, Yuhui Ma, Zaiwang Gu, Shenshen Gu, Jianlong Yang, Jun Cheng, and Jiang Liu. Deep learning with skip connection attention for choroid layer segmentation in oct images. In 2020 42nd Annual International Conference of the IEEE Engineering in Medicine & Biology Society (EMBC), pages 1641– 1645. IEEE, 2020.

27. Yosbelkys Martin Paez, Lucy I Mudie, and Prem S Subramanian. Spaceflight associated neuro-ocular syndrome (sans): A systematic review and future directions. Eye and Brain, pages 105–117, 2020.

28. Javier Mazzaferri, Luke Beaton, Gisèle Hounye, Diane N Sayah, and Santiago Costantino. Open-source algorithm for automatic choroid segmentation of oct volume reconstructions. Scientific reports, 7(1):42112, 2017.

29. Marcel N Menke, Gilbert T Feke, and Clement L Trempe. Oct measurements in patients with optic disc edema. Investigative ophthalmology & visual science, 46(10):3807– 3811, 2005.

30. An L Moens, Hunter C Champion, Marc J Claeys, Barbara Tavazzi, Pawel M Kaminski, Michael S Wolin, Dirk J Borgonjon, Luc Van Nassauw, Azeb Haile, Muz Zviman, et al. High-dose folic acid pretreatment blunts cardiac dysfunction during ischemia coupled to maintenance of high-energy phosphates and reduces postreperfusion injury. Circulation, 117(14):1810–1819, 2008.

31. An L Moens, Christiaan J Vrints, Marc J Claeys, Jean-Pierre Timmermans, Hunter C Champion, and David A Kass. Mechanisms and potential therapeutic targets for folic acid in cardiovascular disease. American Journal of Physiology-Heart and Circulatory Physiology, 294(5):H1971–H1977, 2008.

32. Sarah Mrejen and Richard F Spaide. Optical coherence tomography: imaging of the choroid and beyond. Survey of ophthalmology, 58(5):387–429, 2013.

33. AM Nguyen, T Balmitgere, M Bernard, C Tilikete, A Vighetto, et al. Detection of mild papilloedema using spectral domain optical coherence tomography. British journal of ophthalmology, 96(3):375–379, 2012.

34. Joshua Ong, Andrew G Lee, and Heather E Moss. Head-down tilt bed rest studies as a terrestrial analog for spaceflight associated neuro-ocular syndrome. Frontiers in Neurology, 12:648958, 2021.

35. Joshua Ong, William Tarver, Tyson Brunstetter, Thomas Henry Mader, C Robert Gibson, Sara S Mason, and Andrew Lee. Spaceflight associated neuro-ocular syndrome: proposed pathogenesis, terrestrial analogues, and emerging countermeasures. British Journal of Ophthalmology, 107(7):895–900, 2023.

36. Laura P Pardon, Han Cheng, Pratik Chettry, and Nimesh B Patel. Optic nerve head morphological changes over 12 hours in seated and head-down tilt postures. Investigative Ophthalmology & Visual Science, 61(13):21–21, 2020.

37. Arshi Parvaiz, Muhammad Anwaar Khalid, Rukhsana Zafar, Huma Ameer, Muhammad Ali, and Muhammad Moazam Fraz. Vision transformers in medical computer vision—a contemplative retrospection. Engineering Applications of Artificial Intelligence, 122: 106126, 2023.

38. Zarana S Patel, Tyson J Brunstetter, William J Tarver, Alexandra M Whitmire, Sara R Zwart, Scott M Smith, and Janice L Huff. Red risks for a journey to the red planet: The highest priority human health risks for a mission to mars. npj Microgravity, 6(1):33, 2020.

39. Diane N Sayah, Denise Descovich, Santiago Costantino, and Mark R Lesk. The association between the pulsatile choroidal volume change and ocular rigidity. Ophthalmology Science, page 100576, 2024.

40. Ari Shinojima, Ken-ichi Iwasaki, Ken Aoki, Yojiro Ogawa, Ryo Yanagida, and Mitsuko Yuzawa. Subfoveal choroidal thickness and foveal retinal thickness during head-down tilt. Aviation, space, and environmental medicine, 83(4):388–393, 2012.

41. Patrick A Sibony, Steven S Laurie, Connor R Ferguson, Laura P Pardon, Millennia Young, F James Rohlf, and Brandon R Macias. Ocular deformations in spaceflight-associated neuro-ocular syndrome and idiopathic intracranial hypertension. Investigative Ophthalmology & Visual Science, 64(3):32–32, 2023.

42. Sumit Randhir Singh, Kiran Kumar Vupparaboina, Abhilash Goud, Kunal K Dansingani, and Jay Chhablani. Choroidal imaging biomarkers. Survey of Ophthalmology, 64(3): 312–333, 2019.

43. Marissé Masís Solano, Emmanuelle Richer, Farida Cheriet, Mark R Lesk, and Santiago Costantino. Mapping pulsatile optic nerve head deformation using oct. Ophthalmology Science, 2(4):100205, 2022.

44. Marissé Masís Solano, Charles Bélanger Nzakimuena, Rémy Dumas, Mark R Lesk, and Santiago Costantino. Ocular rigidity and choroidal thickness changes in response to microgravity: A case study. American Journal of Ophthalmology Case Reports, 32: 101940, 2023.

45. Marissé Masís Solano, Remy Dumas, Mark R Lesk, and Santiago Costantino. Ocular biomechanical responses to long-duration spaceflight. IEEE Open Journal of Engineering in Medicine and Biology, 2024.

46. Xiaodan Sui, Yuanjie Zheng, Benzheng Wei, Hongsheng Bi, Jianfeng Wu, Xuemei Pan, Yilong Yin, and Shaoting Zhang. Choroid segmentation from optical coherence tomography with graph-edge weights learned from deep convolutional neural networks. Neurocomputing, 237:332–341, 2017.

47. Hans Thisanke, Chamli Deshan, Kavindu Chamith, Sachith Seneviratne, Rajith Vidanaarachchi, and Damayanthi Herath. Semantic segmentation using vision transformers: A survey. Engineering Applications of Artificial Intelligence, 126:106669, 2023.

48. Ethan Waisberg, Joshua Ong, Mouayad Masalkhi, Kazuhito Shimada, and Andrew G Lee. Artificial gravity as a potential countermeasure for spaceflight associated neuroocular syndrome. Eye, pages 1–2, 2024.

49. Yaqi Wang, Zehua Yang, Xindi Liu, Zhi Li, Chengyu Wu, Yizhen Wang, Kai Jin, Dechao Chen, Gangyong Jia, Xiaodiao Chen, et al. Pgkd-net: Prior-guided and knowledge diffusive network for choroid segmentation. Artificial Intelligence in Medicine, 150:102837, 2024.

50. Stephen M Wilson, Alexa Bautista, Melodie Yen, Stefanie Lauderdale, and Dana K Eriksson. Validity and reliability of four language mapping paradigms. NeuroImage: Clinical, 16:399–408, 2017.

51. Peter Wostyn, Charles R Gibson, and Thomas H Mader. Optic disc edema in astronauts from a choroidal point of view. Aerospace Medicine and Human Performance, 93(4): 396–398, 2022.

52. L Wu, M Masis, and E Hernandez-Bogantes. Choroidal imaging with spectral-domain optical coherence tomography, enhanced depth imaging may lead to a broader understanding of the pathogenesis of several eye diseases. Retina Surgery Global Perspectives, 114:39–42, 2011.

53. Wenjun Wu, Yan Gong, Huaying Hao, Jiong Zhang, Pan Su, Qifeng Yan, Yuhui Ma, and Yitian Zhao. Choroidal layer segmentation in oct images by a boundary enhancement network. Frontiers in Cell and Developmental Biology, 10:1060241, 2022.

54. Enze Xie, Wenhai Wang, Zhiding Yu, Anima Anandkumar, Jose M Alvarez, and Ping Luo. Segformer: Simple and efficient design for semantic segmentation with transformers. Advances in neural information processing systems, 34:12077–12090, 2021.

55. Meng Xuan, Wei Wang, Danli Shi, James Tong, Zhuoting Zhu, Yu Jiang, Zongyuan Ge, Jian Zhang, Gabriella Bulloch, Guankai Peng, et al. A deep learning–based fully automated program for choroidal structure analysis within the region of interest in myopic children. Translational Vision Science & Technology, 12(3):22–22, 2023.

56. Haoran Zhang, Jianlong Yang, Ce Zheng, Shiqing Zhao, and Aili Zhang. Annotation-efficient learning for oct segmentation. Biomedical Optics Express, 14(7):3294–3307, 2023.

57. Sara R Zwart, C Robert Gibson, Thomas H Mader, Karen Ericson, Robert Ploutz-Snyder, Martina Heer, and Scott M Smith. Vision changes after spaceflight are related to alterations in folate-and vitamin b-12-dependent one-carbon metabolism. The Journal of nutrition, 142(3):427–431, 2012.

58. Sara R Zwart, Jesse F Gregory, Steven H Zeisel, Charles R Gibson, Thomas H Mader, Jason M Kinchen, Per M Ueland, Robert Ploutz-Snyder, Martina A Heer, and Scott M Smith. Genotype, b-vitamin status, and androgens affect spaceflight-induced ophthalmic changes. The FASEB Journal, 30(1):141, 2016.

59. Sara R Zwart, Charles R Gibson, Jesse F Gregory, Thomas H Mader, Patrick J Stover, Steven H Zeisel, and Scott M Smith. Astronaut ophthalmic syndrome. FASEB Journal, 31(9):3746–3756, 2017.

