## Supplementary Information for "Spatial and temporal changes in choroid morphology associated with long-duration spaceflight"

#### A. Datasets summary

For OCT videos, there was an average interval between return from spaceflight and postflight acquisition of 11.5 ± 10.2 days (the minimum interval was 3 days, and the maximum was 35 days).

For OCT videos, all acquisitions were obtained between 8h00 and 16h00, with a mean daytime of 11h08 ± 2.2 hr. There was a range of differences between the acquisition time of pre- and post-spaceflight acquisition sessions of 0-8 hr, and a mean difference of 2.8 ± 2.1 hr. For OCT volumes, all acquisitions were obtained between 13h17 and 20h00, with a mean daytime of 16h50 ± 1.7 hr. There was a range of differences between the acquisition time of initial and subsequent timepoints of 0-5.857 hr, and a mean difference of 2.7 ± 1.7 hr.

A datasets summary table is provided below.

**Table S1. Datasets summary table.** The table provides characteristics of OCT videos and volumes.

| | dataset 1 | dataset 2 |
|---|---|---|
| dataset name | space macular videos | space macular volumes |
| subjects | 13 astronauts | 6 astronauts |
| sex | 9 males, 4 females | 5 males, 1 female |
| age [yo ± SD] | 46.8 ± 8.9 | 47.5 ± 9 |
| timepoints (L launch, R return, m month, d day) | 2 (pre-, post-spaceflight) | 4-6 (L-21 to 18 m, L-9 to 6 m, L+30 d, L+90 d, R-30 d, R+1 to 3 d) |
| eyes | OU | OU |
| # scans/eye [videos OR volumes] | 1-5 videos | 1-2 volumes |
| scan size [px × px] | 768 × 496 | 512 × 496 |
| approximate horizontal resolution [µm/px] | 11 | 11 |
| approximate vertical resolution [µm/px] | 3.9 | 3.9 |
| approximate physical area [mm × mm] | 8.4 × 1.9 | 5.6 × 1.9 |
| # frames per scan [B-scans] | 441 (average) | 97 or 193 |
| # frames total for the dataset | 45,439 | 11,719 |

#### B. Data pipeline flowcharts

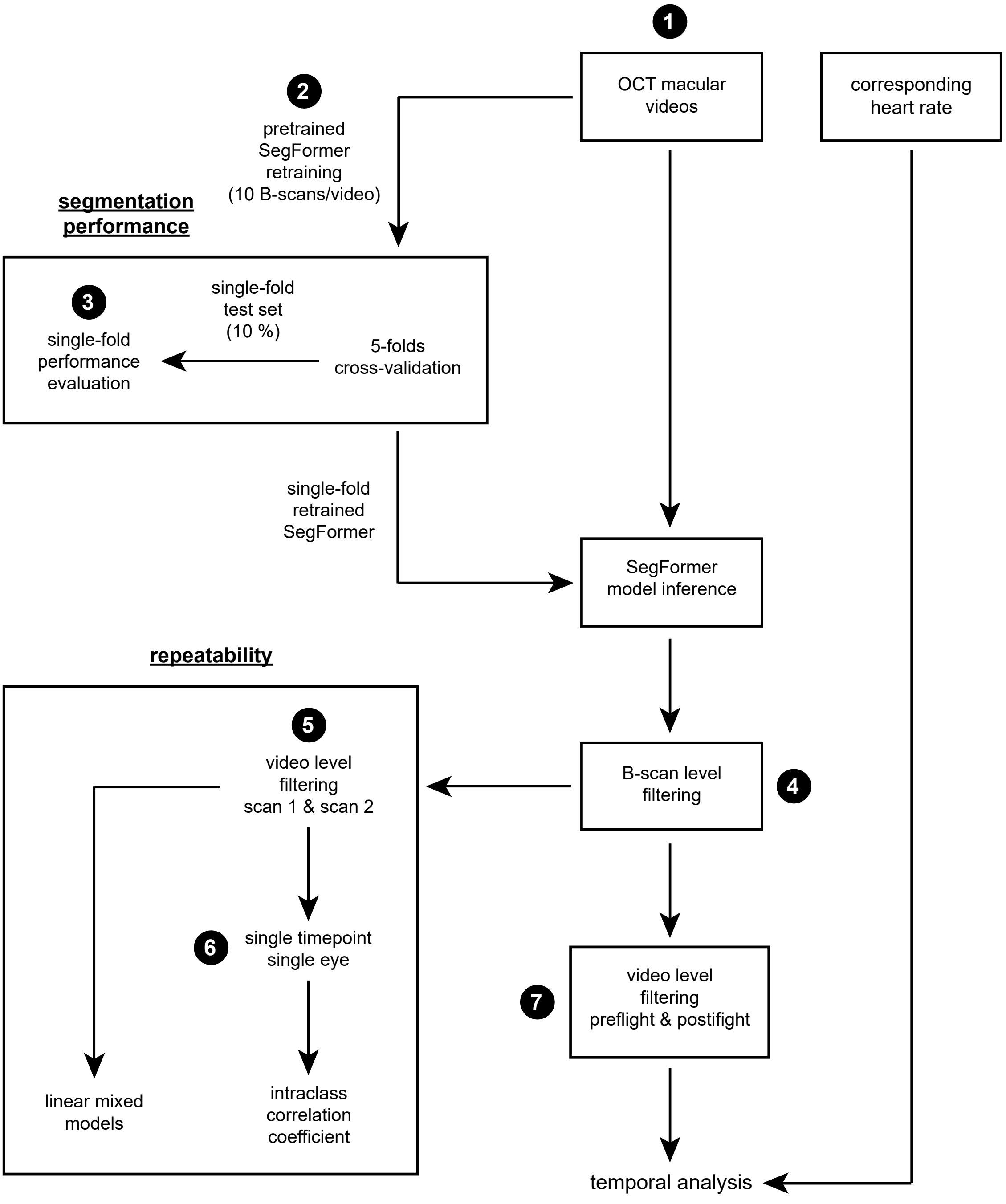


**Figure S1. Flowchart of OCT videos data pipeline.** Numbered circles indicate steps where the dataset is altered from initial conditions (number of subjects, eyes, videos and B-scans).

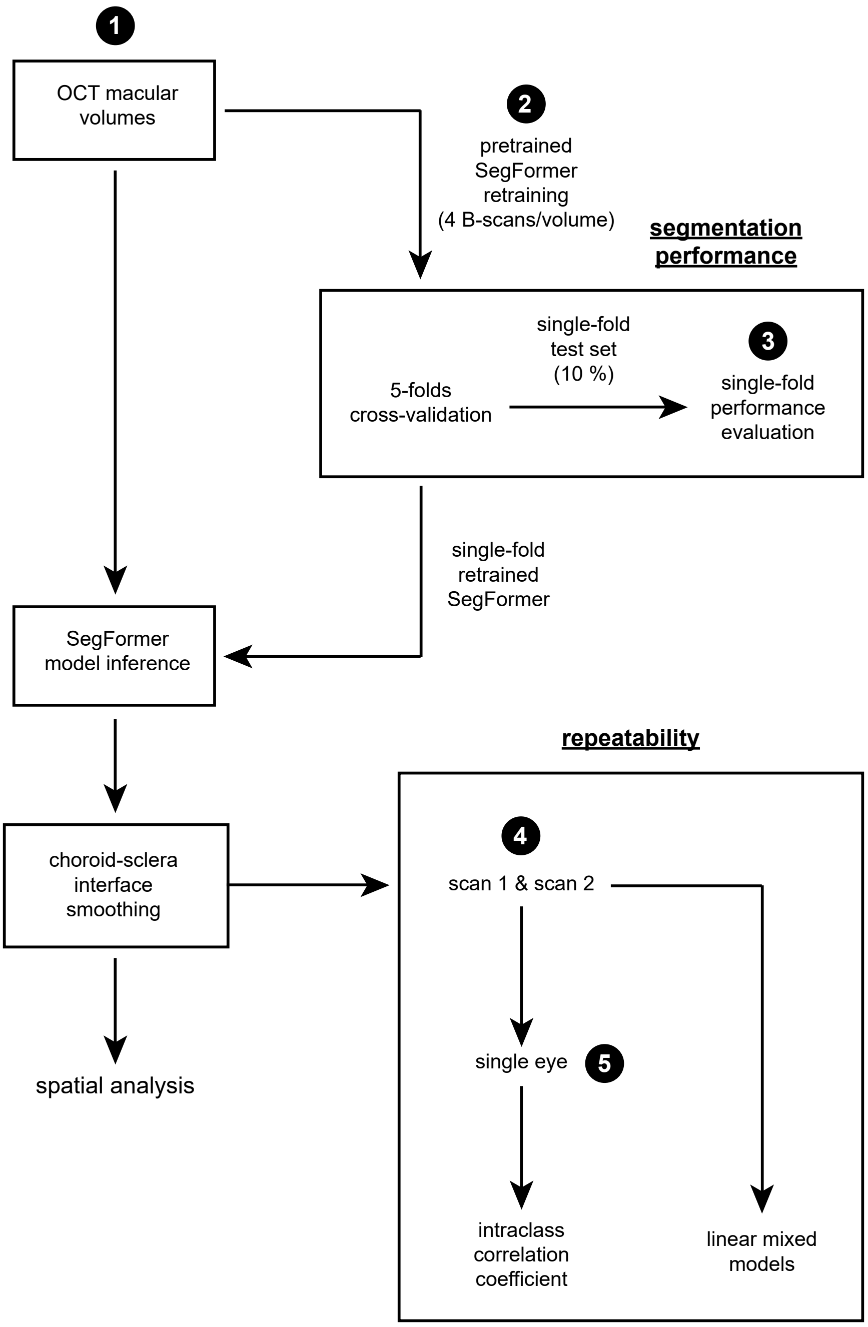


**Figure S2. Flowchart of OCT volumes data pipeline.** Similarly as for videos, numbered circles indicate dataset alteration (number of subjects, eyes, volumes and B-scans).

##### C. Post-processing

Post-processing of the deep learning choroid segmentation output mask first consists of applying an opening operation to minimize undesirable false positive regions. Opening is performed for 5 iterations using a 5 pixels width × 3 pixels height rectangular kernel. To accurately identify Bruch's membrane (BM) and the choroid-sclera interface (CSI) as the upper and lower choroid boundaries, respectively, including when several isolated false positive regions are present, the binary image undergoes a series of boundaries extraction steps:

The first boundaries extraction step consists of iterating through each of the mask's objects to obtain images of the BM and CSI candidate segments. For each object, the contour is obtained and any void inside the region is filled. An argmax function is applied in the vertical axis direction to obtain locations of the BM candidate segments. BM candidate segments are then combined into a single binary image. Any candidate pixel at the top edge of the BM candidate segments image is removed, and candidate segments less than 10 pixels in size are discarded. Similar processing is performed to obtain the CSI candidate segments image, removing any candidate pixels at the bottom image edge instead of at the top edge.

The second boundaries extraction step consists of connecting boundary paths for both the BM and CSI candidate segments images. In both cases, an iteration through all candidate segments is performed to identify right and left segment extremities and their coordinates. Each extremity is connected to its closest opposite side extremity, or to the opposite edge of the image where crossing with an existing segment does not occur. Once extremities are connected, only the largest connected region in the boundary candidate segments image is kept. Then, the extremities connections process is performed once more to ensure the remaining region connects with the left and right edges of the image.

For the single remaining region in each boundary candidate segments image, the shortest path between the left and right edge of the image is determined and all other pixels are discarded. To prevent the BM and CSI from crossing each other, if any BM location is found at a lower location than the corresponding CSI location along the vertical axis of both boundary candidate segments images, then the BM location is changed to one pixel above the corresponding CSI location. The BM and CSI are each returned as arrays of locations along the vertical axis of their respective candidate segments images.

**Algorithm 1** Side points iteration (e. g. right side, $P_2$)

```
for every point Pi do
    gather all left-side extremities on right side of current extremity (P2)
    opposite candidate points array, Ao = [7, 9, 11, 13, 15, 17]
    gather all right-side extremities on right side of current extremity (P2)
    edge most points array, Ae = [6, 4, 8, 10, 12, 14, 16, 18]
    if Ao is not empty then
        obtain list of distances from current point to opposite side candidate points
        distances array, Ad = [10, 34, 52, 41, 58, 62]
        while Ad not empty do
            gather point coordinate corresponding to minimum distance in Ad (P7)
            connect minimum distance point (P7) to current extremity (P2)
            if no line crossing
            and line vertical distance < than fraction of image height
            and line horizontal distance < than fraction of image width then
                draw line in the image
                end while loop
            else
                remove current distance from Ad
                continue while loop
            end if
        end while [Ad not empty]
    else if Ao is empty
    and Ae is empty then
        current extremity is edge most,
        connect current extremity directly horizontally to the same side image edge
    end if
end for
```

**Figure S3. Side points iteration algorithm.** Pseudocode of the side points iteration algorithm.

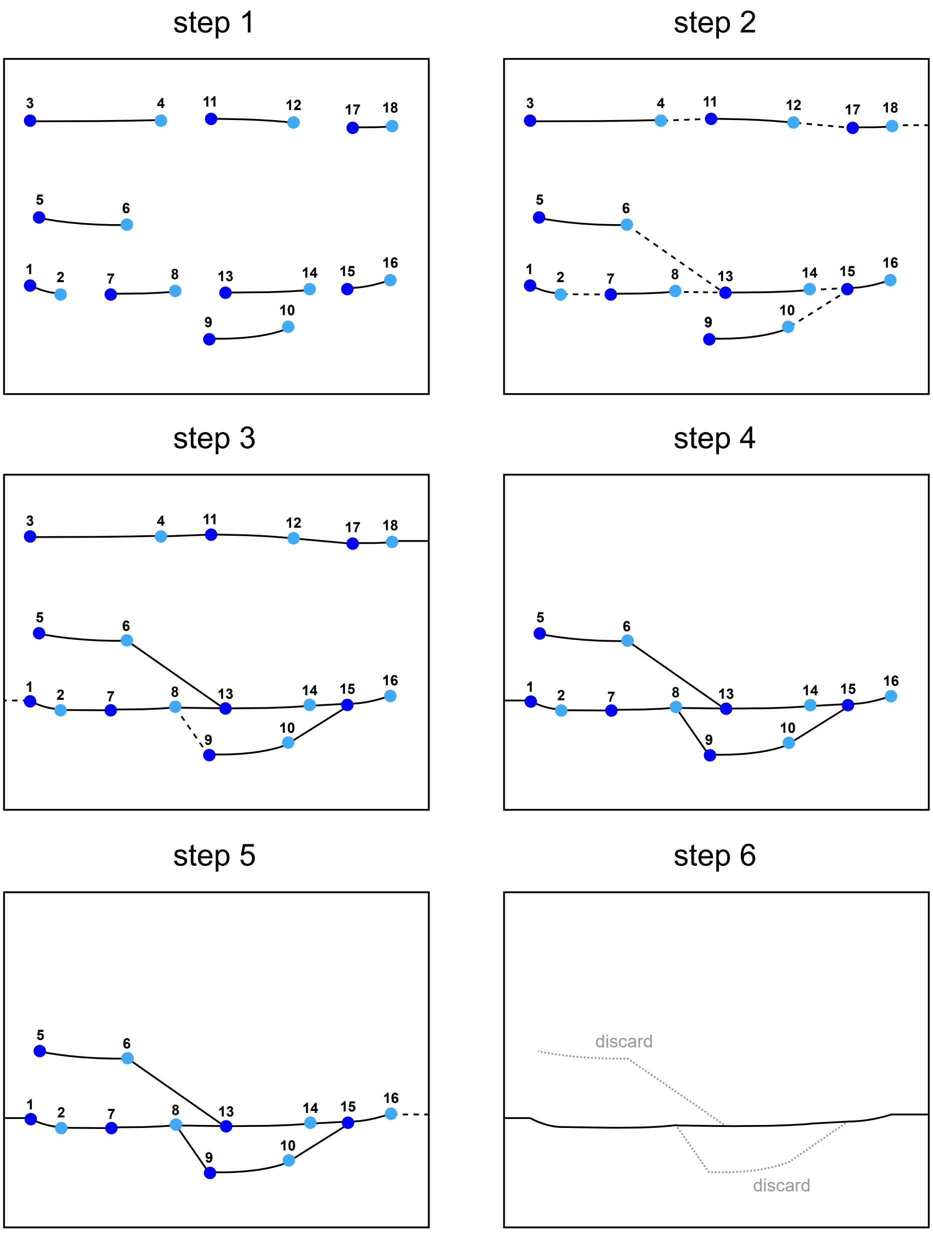


**Figure S4. Illustrated post-processing steps.** (**step 1**) Boundary candidate points identification. (**step 2**) Side points iteration for right side points. (**step 3**) Repetition of side points iteration for left side points. (**step 4**) Only the largest region of interest in the image is kept. (**step 5**) Repetition of side points iteration for right and left sides to ensure image side edges connections. (**step 6**) Only the shortest path between image right and left side edges is kept.

#### D. Space macular videos and volumes choroid segmentation cross-validation and post-processing results

Equations relating to the inaccurate segmentation criterion are provided below

$$CT_{B-scan\ mean} = \frac{\sum_{i=1}^{n} CT_{B-scan\ column,\ i}}{n} \tag{1}$$

$$PC_{CT\ diff} = \frac{|CT_{manual} - CT_{model}|}{CT_{manual}} \times 100\% \tag{2}$$

$$C_{inaccurate} = \begin{cases} 1 & \text{if } PC_{CT\ diff} \geq 20\%, \\ 0 & \text{otherwise.} \end{cases} \tag{3}$$

where $CT_{\text{B-scan column}}$ is choroid thickness corresponding to a column in a B-scan image, $CT_{\text{B-scan mean}}$ is the mean of all column choroid thicknesses in a B-scan, $PC_{\text{CT diff}}$ is the absolute difference between $CT_{\text{B-scan mean}}$ obtained through manual segmentation, $CT_{\text{manual}}$, and $CT_{\text{B-scan mean}}$ obtained through automated segmentation, $CT_{\text{model}}$, as a percentage of $CT_{\text{manual}}$, and $C_{\text{inaccurate}}$ is the inaccurate segmentation criterion.

Equations relating to the segmentation failure criterion are provided below

$$CT_{B-scan\ max} = \max_{1 \leq i \leq n} \{CT_{B-scan\ column,\ i}\} \tag{4}$$

$$PC_{max\ CT\ diff} = \frac{|CT_{B-scan\ max} - CT_{manual}|}{CT_{manual}} \times 100\% \tag{5}$$

$$C_{failure} = \begin{cases} 1 & \text{if } PC_{max\ CT\ diff} \geq 60\% \text{ and } \sigma_{model} \geq 2.5 \cdot \sigma_{manual}, \\ 0 & \text{otherwise.} \end{cases} \tag{6}$$

where $CT_{\text{B-scan max}}$ is the maximum choroid thickness of all column choroid thicknesses in a B-scan, $\sigma_{\text{model}}$ is the standard deviation of a set of $CT_{\text{B-scan column}}$ obtained through automatic segmentation, $\sigma_{\text{manual}}$ is the standard deviation of a set of $CT_{\text{B-scan column}}$ obtained through manual segmentation, $PC_{\text{max CT diff}}$ is the absolute difference between $CT_{\text{B-scan max}}$ and $CT_{\text{manual}}$ as a percentage of $CT_{\text{manual}}$, and $C_{\text{failure}}$ is the segmentation failure criterion.

Figure S5 displays 5-fold cross-validation choroid segmentation performance results for space macular videos and volumes. Figure S6 depicts 5-fold cross validation post-processing results for space macular videos and volumes. As shown on the left side of figure S6a, higher fold-averaged $PC_{\text{max CT diff}}$ values indicate a higher occurrence of failed segmentations in the absence of post-processing. Similarly, the stacked bar plots on the left side of figure S6b indicate that images were more likely to meet the inaccurate segmentation and segmentation failure criteria for a given fold in the absence of post-processing. The same analysis for the macular volumes dataset, as shown in S6d and e, suggests that images in that dataset were less problematic.

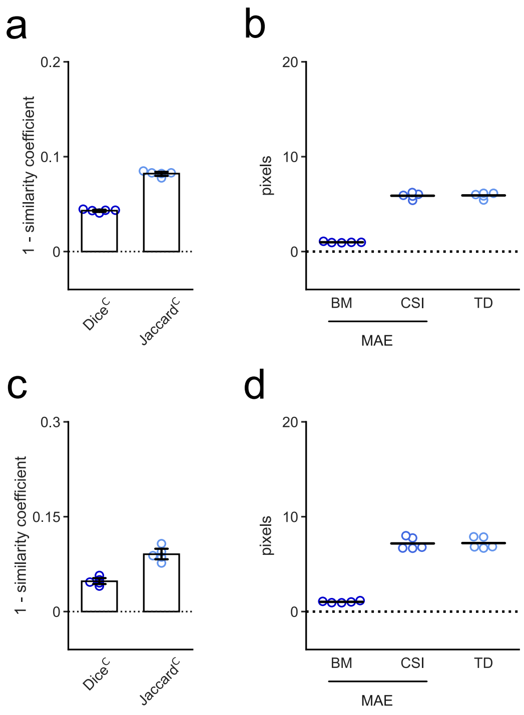


**Figure S5. 5-fold cross-validation choroid segmentation performance results for space macular videos and volumes.** (**a**) Dice$^C$ and Jaccard$^C$ results for the test set of 5-fold cross-validation space macular videos-retrained models. (**b**) BM and CSI layers mean absolute error, and thickness difference for the test set of 5-fold cross-validation space macular videos-retrained models (**c**) Dice$^C$ and Jaccard$^C$ results for the test set of 5-fold cross-validation space macular volumes-retrained models. (**d**) BM and CSI layers mean absolute error, and thickness difference for the test set of 5-fold cross-validation space macular volumes-retrained models.

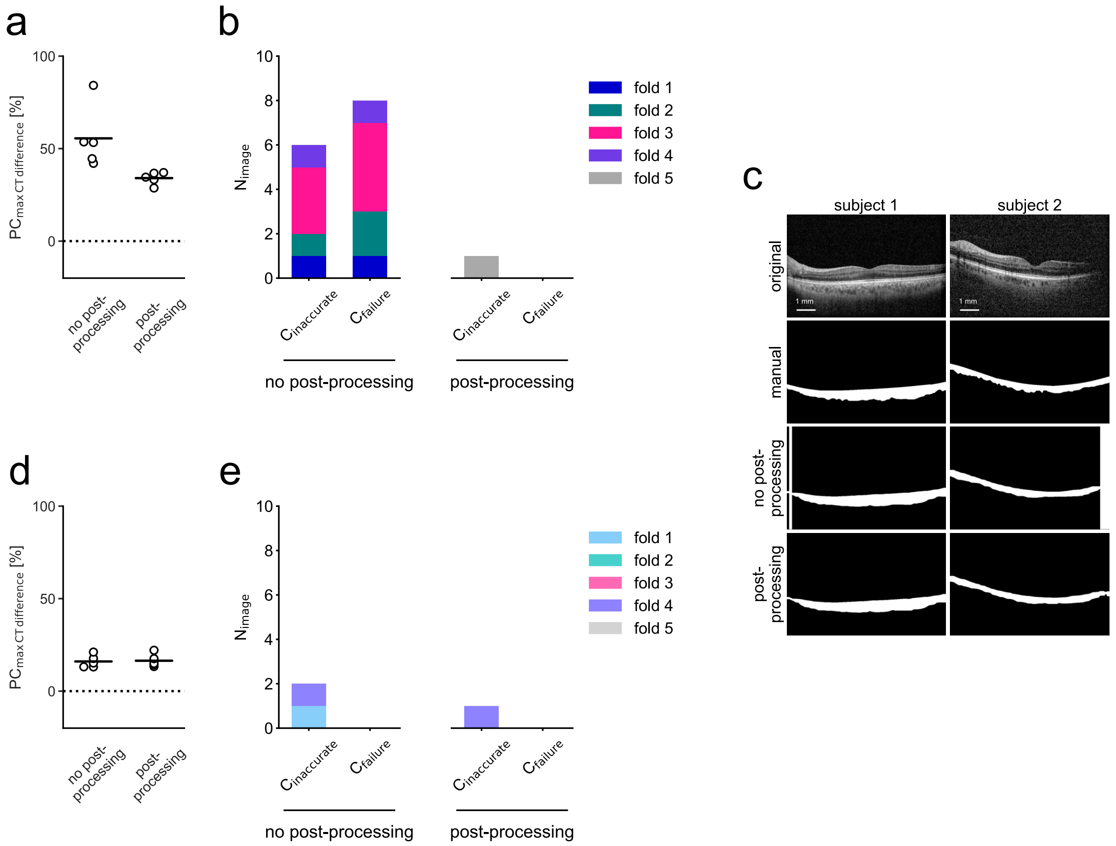


**Figure S6. 5-fold cross-validation post-processing results for space macular videos and volumes.** (**a**) $PC_{max\ CT\ diff}$ 5-fold cross-validation results without and with post-processing for choroids segmented using OCT videos-retrained models. (**b**) Stacked bar charts of inferred choroid segmentations (left) without and (right) with post-processing meeting inaccurate segmentation and segmentation failure criteria for 5-fold cross-validation OCT videos-retrained models. (**c**) Original B-scans and binary choroid segmentation masks for 2 subjects obtained manually, through model inference without and with post-processing. Binary choroid segmentation masks for which post-processing was not applied show segmentation failures. (**d**) $PC_{max\ CT\ diff}$ 5-fold cross-validation results without and with post-processing for choroids segmented using OCT volumes-retrained models. (**e**) Stacked bar charts of inferred choroid segmentations (left) without and (right) with post-processing meeting inaccurate segmentation and segmentation failure criteria for 5-fold cross-validation OCT volumes-retrained models.

#### E. Luminal and stromal regions processing

Individual B-scan processing to obtain a binary mask of luminal area and towards performing CVI calculation were implemented similarly as described in literature[1; 4; 2]. First, Niblack local thresholding was applied in Python (v3.10.0) through sickit-image (0.19.2). The Niblack function was implemented with a specified window of 35 pixels and a constant value, k, of 0.8. Following Niblack local thresholding, dilation and closing morphological operations, both using a 3 × 3 pixels kernel, were applied to the image. The final binarized luminal area image was achieved by keeping only the processed region corresponding to the choroid layer. The CVI for individual B-scans is obtained through the following equation

$$CVI = \frac{LA}{CA} \tag{7}$$

where LA is luminal area and CA is total choroid area. Stromal area is related to luminal area through the following equation

$$SA = CA - LA \tag{8}$$

where SA is stromal area. The CVI for OCT volumes is obtained through the following equation

$$CVI = \frac{LT}{CT} \tag{9}$$

where LT is luminal thickness averaged over the specified macular region, and CT is choroid thickness averaged over the specified macular region. Stromal thickness is related to luminal thickness through the following equation

$$ST = CT - LT \tag{10}$$

where ST is stromal thickness.

#### F. ETDRS subfields

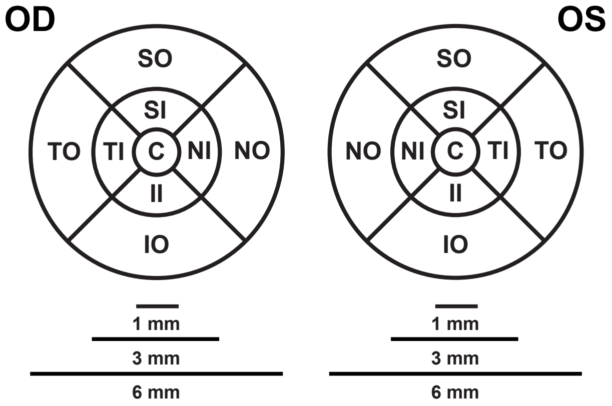


**Figure S7. ETDRS subfields.** Nine ETDRS subfields for right eye (OD) and left eye (OS). (C center, SI superior inner, NI nasal inner, II inferior inner, TI temporal inner, SO superior outer, NO nasal outer, IO inferior outer, TO temporal outer).

#### G. Transforms implementation details

The Lomb–Scargle periodogram and Lomb–Scargle spectrogram were implemented in Python (v3.10.0) through SciPy (1.8.0). At an automatic real time frame averaging of 5 frames, the OCT device used had an approximate acquisition rate of 21.662 frames/second (0.0462 seconds period). For the spectrogram, a 40 elements window ($\sim$1.848 seconds) was used with a 2 elements step ($\sim$0.0924 seconds).

As shown in figure S8, peaks at the specified sinusoid frequency are visible in the Lomb–Scargle periodogram. For the Lomb–Scargle spectrogram, the peaks are sustained over the time interval.

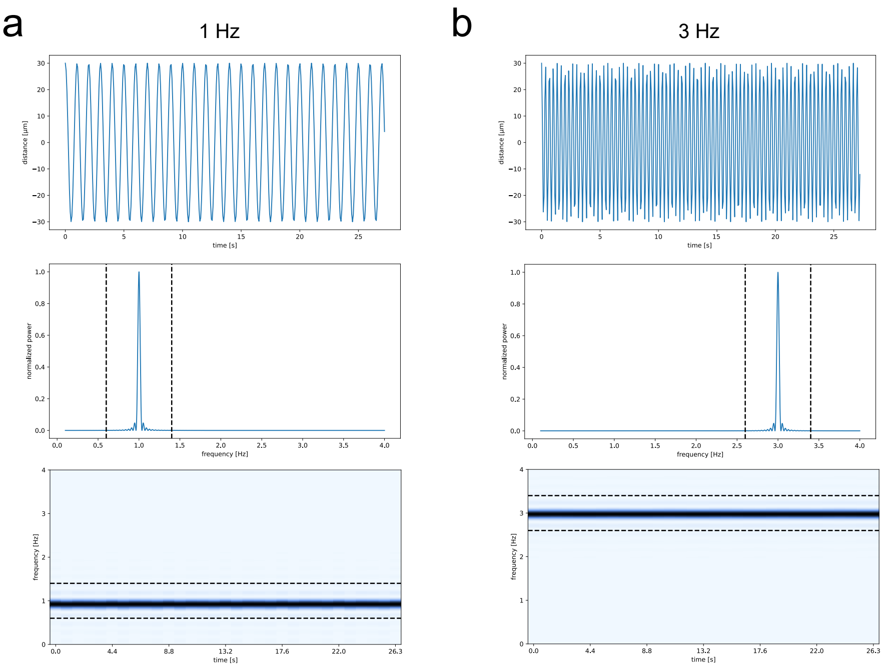


**Figure S8. Transforms calibration.** (**a**) Transforms calibration with a 1 Hz sinusoid. Top row features a sinusoidal timeseries, second row features the Lomb–Scargle periodogram and third row features the Lomb–Scargle spectrogram. Dashed line on the Lomb–Scargle periodogram and Lomb–Scargle spectrogram show values $\pm$ 40 % of the specified frequency value. (**b**) Transforms calibration with a 3 Hz sinusoid.

#### H. Scheme for volumes manual labelling

| | OD volume | 1 | 2 | 3 | 4 | 5 | 6 | 7 | 8 | 9 | 10 | 11 | 12 | 13 | 14 | 15 | 16 | 17 | 18 | 19 | 20 |
|---|---|---|---|---|---|---|---|---|---|---|---|---|---|---|---|---|---|---|---|---|---|
| L - 21 to 18 m | 1 | x | x | | | | | | x | x | | | | | | | | | | | |
| L - 21 to 18 m | 2 | | x | x | | | | | | x | x | | | | | | | | | | |
| L - 9 to 6 m | 1 | | | x | x | | | | | | x | x | | | | | | | | | |
| L - 9 to 6 m | 2 | | | | x | x | | | | | | x | x | | | | | | | | |
| L + 30 d | 1 | | | | | x | x | | | | | | x | x | | | | | | | |
| L + 30 d | 2 | | | | | | x | x | | | | | | x | x | | | | | | |
| L + 90 d | 1 | | | | | | | x | x | | | | | | x | x | | | | | |
| L + 90 d | 2 | | | | | | | | x | x | | | | | | x | x | | | | |
| R - 30 d | 1 | | | | | | | | | x | x | | | | | | x | x | | | |
| R - 30 d | 2 | | | | | | | | | | x | x | | | | | | x | x | | |
| R + 1 to 3 d | 1 | | | | | | | | | | | x | x | | | | | | x | x | |
| R + 1 to 3 d | 2 | | | | | | | | | | | | x | x | | | | | | x | x |

| | OS volume | 1 | 2 | 3 | 4 | 5 | 6 | 7 | 8 | 9 | 10 | 11 | 12 | 13 | 14 | 15 | 16 | 17 | 18 | 19 | 20 |
|---|---|---|---|---|---|---|---|---|---|---|---|---|---|---|---|---|---|---|---|---|---|
| L - 21 to 18 m | 1 | x | x | | | | | | x | x | | | | | | | | | | | |
| L - 21 to 18 m | 2 | | x | x | | | | | | x | x | | | | | | | | | | |
| L - 9 to 6 m | 1 | | | x | x | | | | | | x | x | | | | | | | | | |
| L - 9 to 6 m | 2 | | | | x | x | | | | | | x | x | | | | | | | | |
| L + 30 d | 1 | | | | | x | x | | | | | | x | x | | | | | | | |
| L + 30 d | 2 | | | | | | x | x | | | | | | x | x | | | | | | |
| L + 90 d | 1 | | | | | | | x | x | | | | | | x | x | | | | | |
| L + 90 d | 2 | | | | | | | | x | x | | | | | | x | x | | | | |
| R - 30 d | 1 | | | | | | | | | x | x | | | | | | x | x | | | |
| R - 30 d | 2 | | | | | | | | | | x | x | | | | | | x | x | | |
| R + 1 to 3 d | 1 | | | | | | | | | | | x | x | | | | | | x | x | |
| R + 1 to 3 d | 2 | | | | | | | | | | | | x | x | | | | | | x | x |

**Figure S9. Scheme for volumes manual labelling.** Dataset 2 volumes manual labelling scheme. (L launch, R return, m months, d days).

#### I. Additional choroid layer segmentation and lumen segmentation performance evaluation

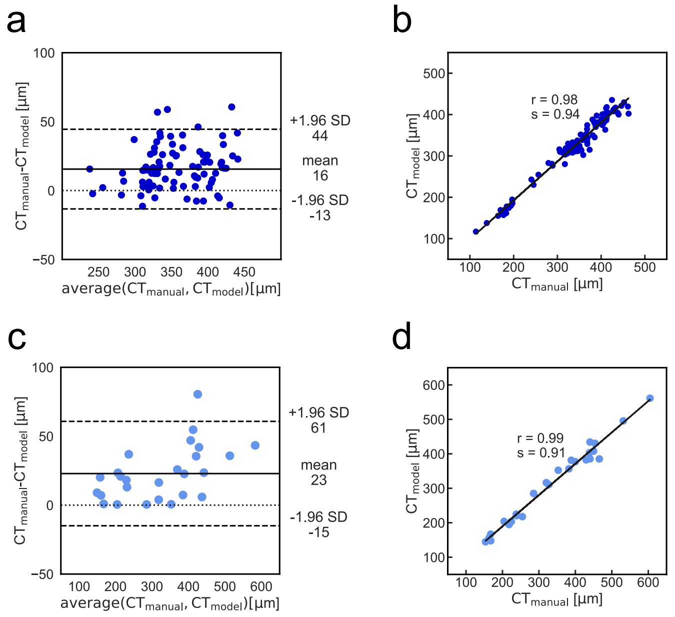


**Figure S10. Choroid layer segmentation additional performance evaluation.** (**a**) Bland-Altman plot illustrating the agreement between CT obtained manually and through model inference for the OCT videos test set. (**b**) Scatterplot and regression line of model inferred CT against manually obtained CT for the OCT videos test set. r Pearson correlation coefficient, s regression line slope. (**c**) Bland-Altman plot illustrating the agreement between CT obtained manually and through model inference for the OCT volumes test set. (**d**) Scatterplot and regression line of model inferred CT against manually obtained CT for the OCT volumes test set. r Pearson correlation coefficient, s regression line slope.

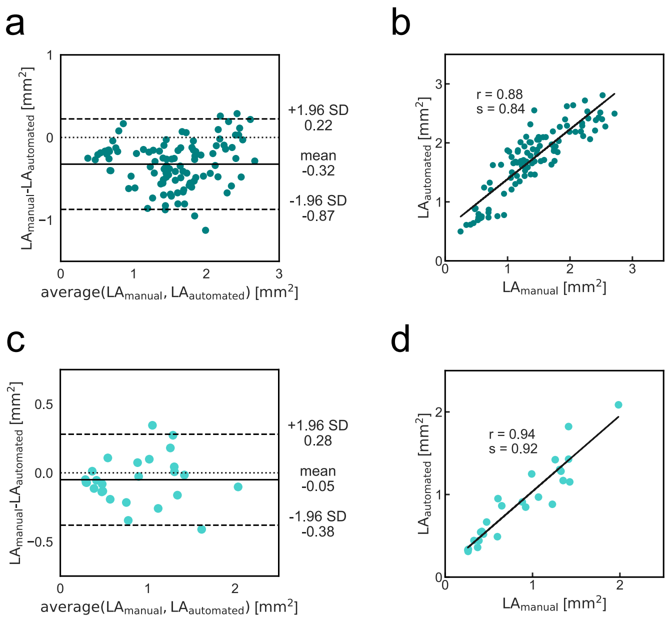


**Figure S11. Choroid lumen segmentation additional performance evaluation.** (**a**) Bland-Altman plot illustrating the agreement between LA obtained manually and LA obtained automatically for the OCT videos test set. (**b**) Scatterplot and regression line of automatically obtained LA against manually obtained LA for the OCT videos test set. r Pearson correlation coefficient, s regression line slope. (**c**) Bland-Altman plot illustrating the agreement between LA obtained manually and LA obtained automatically for the OCT volumes test set. (**d**) Scatterplot and regression line of automatically obtained LA against manually obtained LA for the OCT volumes test set. r Pearson correlation coefficient, s regression line slope.

#### J. Videos repeatability

Repeatability was evaluated for OCT videos by gathering significance from the difference in measurement values between repeated and original scans using linear mixed models. For significance, the first repeated scan from astronaut macular OCT videos including all available eyes from both pre- and post-spaceflight timepoints was used for comparison against the original scan ($n_{sessions}$ = 32, $n_{eyes}$ = 22, $n_{subjects}$ = 13). Repeatability was also evaluated for OCT videos by obtaining the intraclass correlation coefficient (two-way mixed effects, absolute agreement, and single measurement), and the standard error of measurement between measurement values of repeated and original scans. For the latter metrics, only a single eye per astronaut from a single timepoint was used to avoid intra-subject codependence ($n_{sessions}$, $n_{eyes}$, $n_{subjects}$ = 13). The intraclass correlation coefficient computation was implemented in Python (v3.10.0) through pingouin (0.5.2), and the standard error of measurement was computed as described in literature[3].

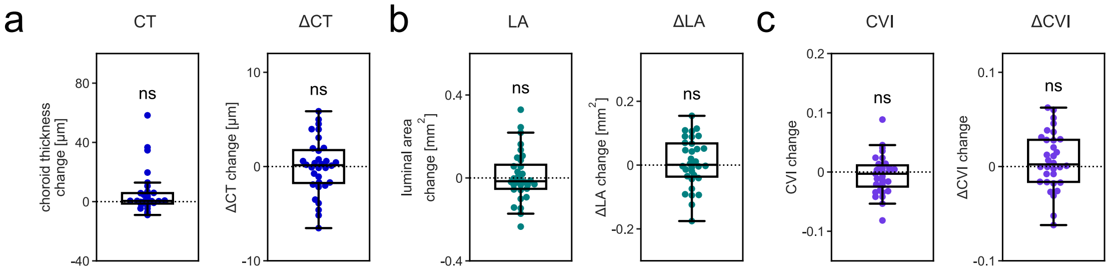


**Figure S12. Temporal OCT choroid pulsatile changes over repeated OCT videos.** (**a**) Difference box plot between repeated and original scan CT and ΔCT measurements obtained over the selected segments for all available astronaut macular OCT videos. (**b**) Difference box plot between repeated and original scan LA and ΔLA measurements. (**c**) Difference box plot between repeated and original scan CVI and ΔCVI measurements. ns nonsignificant, linear mixed model.

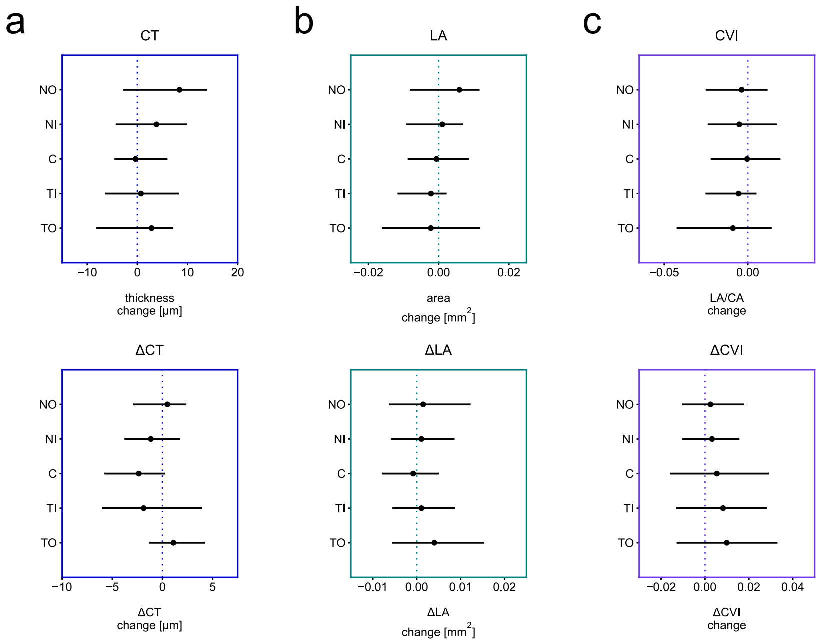


**Figure S13. Temporal OCT choroid pulsatile changes over repeated OCT videos for cross-sectional ETDRS subfields.** Difference plot lines represent the difference interquartile range and dots show the mean difference. (**a**) Difference plot between repeated and original scan CT and ΔCT measurements obtained over the selected segments for all available astronaut macular OCT videos. (**b**) Difference plot between repeated and original scan LA and ΔLA measurements. (**c**) Difference plot between repeated and original scan CVI and ΔCVI measurements. No significant differences were found for any measurements using linear mixed models.

**Table S2. OCT videos significance table (scan 1 — scan 2).** Significance values for OCT videos measures comparisons between repeated and original scans.

scan 1 — scan 2

| | | p-value | $\beta_0$ | $\beta_1$ | 2.5 % lower CI bound | 97.5 % upper CI bound |
|---|---|---|---|---|---|---|
| CT | global | 2.0562E-01 | 303.9085 | 22.8732 | -2.8160 | 13.2302 |
| | NO | 1.2811E-01 | 273.2333 | 23.7594 | -2.3186 | 19.1220 |
| | NI | 5.6067E-01 | 361.2044 | 29.5977 | -9.0669 | 16.6927 |
| | C | 9.5403E-01 | 390.0828 | 31.8877 | -13.1935 | 12.4432 |
| | TI | 9.0511E-01 | 378.4798 | 31.5440 | -11.0241 | 12.4447 |
| | TO | 7.3406E-01 | 330.9792 | 26.9531 | -13.5456 | 19.1943 |
| ΔCT | global | 9.9066E-01 | 14.6193 | 1.2136 | -1.0609 | 1.0483 |
| | NO | 7.5790E-01 | 17.9040 | 2.0570 | -2.6983 | 3.7035 |
| | NI | 5.2273E-01 | 26.6629 | 3.0582 | -4.7176 | 2.4002 |
| | C | 1.3995E-01 | 30.0534 | 3.4580 | -5.4577 | 0.7449 |
| | TI | 3.1130E-01 | 28.8906 | 3.1055 | -5.5143 | 1.7535 |
| | TO | 2.4676E-01 | 22.8613 | 2.9926 | -0.7505 | 2.9363 |
| LA | global | 7.8173E-01 | 1.7036 | 0.1562 | -0.0537 | 0.0713 |
| | NO | 3.6945E-01 | 0.2482 | 0.0279 | -0.0070 | 0.0188 |
| | NI | 8.7497E-01 | 0.2447 | 0.0255 | -0.0118 | 0.0139 |
| | C | 9.1664E-01 | 0.2662 | 0.0284 | -0.0127 | 0.0114 |
| | TI | 7.1187E-01 | 0.2586 | 0.0273 | -0.0138 | 0.0094 |
| | TO | 8.4287E-01 | 0.3134 | 0.0301 | -0.0243 | 0.0199 |
| ΔLA | global | 6.9371E-01 | 0.2656 | 0.0325 | -0.0328 | 0.0493 |
| | NO | 6.9061E-01 | 0.0451 | 0.0066 | -0.0058 | 0.0088 |
| | NI | 6.2937E-01 | 0.0408 | 0.0044 | -0.0033 | 0.0054 |
| | C | 6.8088E-01 | 0.0461 | 0.0047 | -0.0047 | 0.0031 |
| | TI | 7.2708E-01 | 0.0449 | 0.0044 | -0.0050 | 0.0071 |
| | TO | 3.4516E-01 | 0.0607 | 0.0064 | -0.0043 | 0.0123 |
| CVI | global | 5.2853E-01 | 0.6173 | 0.0177 | -0.0186 | 0.0096 |
| | NO | 6.0021E-01 | 0.5866 | 0.0187 | -0.0175 | 0.0101 |
| | NI | 5.8546E-01 | 0.6539 | 0.0229 | -0.0233 | 0.0132 |
| | C | 9.6101E-01 | 0.6559 | 0.0261 | -0.0169 | 0.0161 |
| | TI | 5.6934E-01 | 0.6597 | 0.0245 | -0.0247 | 0.0136 |
| | TO | 4.1429E-01 | 0.6182 | 0.0167 | -0.0303 | 0.0125 |
| ΔCVI | global | 4.5938E-01 | 0.1015 | 0.0135 | -0.0101 | 0.0224 |
| | NO | 7.7317E-01 | 0.1046 | 0.0149 | -0.0147 | 0.0198 |
| | NI | 7.1742E-01 | 0.1058 | 0.0154 | -0.0142 | 0.0206 |
| | C | 4.9819E-01 | 0.0999 | 0.0129 | -0.0103 | 0.0211 |
| | TI | 3.2897E-01 | 0.1009 | 0.0121 | -0.0083 | 0.0247 |
| | TO | 2.3894E-01 | 0.1110 | 0.0153 | -0.0065 | 0.0263 |

**Table S3. OCT videos repeatability metrics table (scan 1 — scan 2).** Intraclass correlation coefficients (ICC) and standard error of measurement (SEM) for OCT videos measures comparisons between repeated and original scans.

scan 1 — scan 2

| | | ICC | SEM |
|---|---|---|---|
| CT | global | 0.9980 | 3.9958 |
| | NO | 0.9876 | 11.2622 |
| | NI | 0.9873 | 13.6652 |
| | C | 0.9862 | 14.4131 |
| | TI | 0.9870 | 13.4013 |
| | TO | 0.9810 | 14.1948 |
| ΔCT | global | 0.8492 | 1.7860 |
| | NO | 0.6564 | 4.7942 |
| | NI | 0.7490 | 7.0315 |
| | C | 0.8170 | 6.9360 |
| | TI | 0.8580 | 5.3141 |
| | TO | 0.8943 | 3.2021 |
| LA | global | 0.9908 | 0.0620 |
| | NO | 0.9899 | 0.0129 |
| | NI | 0.9837 | 0.0138 |
| | C | 0.9899 | 0.0114 |
| | TI | 0.9783 | 0.0155 |
| | TO | 0.9881 | 0.0135 |
| ΔLA | global | 0.5794 | 0.0559 |
| | NO | 0.7214 | 0.0119 |
| | NI | 0.7759 | 0.0065 |
| | C | 0.8301 | 0.0060 |
| | TI | 0.6137 | 0.0084 |
| | TO | 0.6934 | 0.0119 |
| CVI | global | 0.9443 | 0.0187 |
| | NO | 0.9504 | 0.0189 |
| | NI | 0.9252 | 0.0267 |
| | C | 0.9688 | 0.0191 |
| | TI | 0.9264 | 0.0274 |
| | TO | 0.9230 | 0.0227 |
| ΔCVI | global | 0.8053 | 0.0230 |
| | NO | 0.8150 | 0.0226 |
| | NI | 0.8169 | 0.0214 |
| | C | 0.7112 | 0.0217 |
| | TI | 0.7648 | 0.0196 |
| | TO | 0.7809 | 0.0277 |

#### K. Volumes repeatability

Repeatability was evaluated for OCT volumes by gathering significance from the difference in measurement values between repeated and original scans using linear mixed models. For significance, the first repeated scan from astronaut macular OCT volumes including all available eyes from all timepoints was used for comparison against the original scan ($n_{sessions}$, $n_{eyes}$ = 6, $n_{subjects}$ = 3). Repeatability was also evaluated for OCT volumes by obtaining the intraclass correlation coefficient (two-way mixed effects, absolute agreement, and single measurement), and the standard error of measurement between measurement values of repeated and original scans. For the latter metrics, only a single eye per astronaut from a single timepoint was used to avoid intra-subject codependence ($n_{sessions}$, $n_{eyes}$, $n_{subjects}$ = 3). For each OCT volume used to evaluate repeatability, the original scan featured 193 B-scans and the repeated scan featured 97 B-scans.

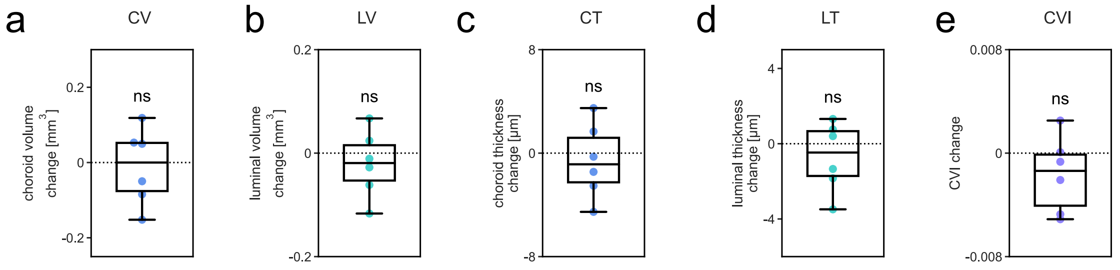


**Figure S14. Spatial OCT choroid changes over repeated OCT volumes.** (**a**) Difference box plot between repeated and original scan CV measurements over the macular region for all available astronaut macular OCT volumes. (**b**) Difference box plot between repeated and original scan LV measurements. (**c**) Difference box plot between repeated and original scan CT measurements averaged over the macular region. (**d**) Difference box plot between repeated and original scan LT measurements. (**e**) Difference box plot between repeated and original scan CVI measurements. ns nonsignificant, linear mixed model.

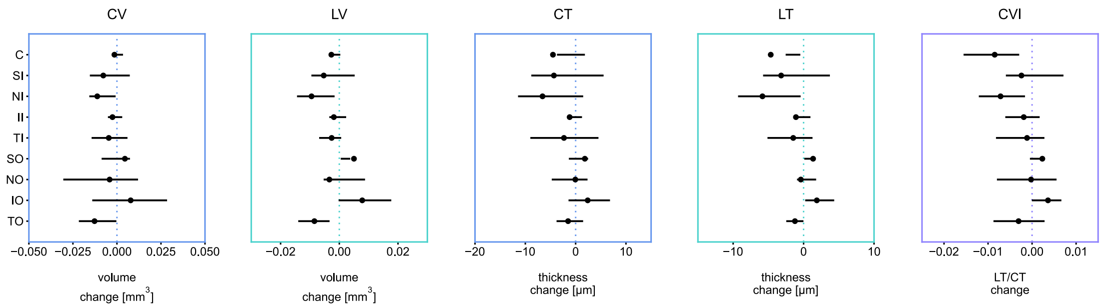


**Figure S15. Spatial OCT choroid changes over repeated OCT volumes.** Difference plots between repeated and original scan CV and LV measurements over the specified macular regions, and CT, LT and CVI measurements averaged over the specified macular regions for all available astronaut macular OCT volumes. ETDRS subfield differences are shown. Difference plot lines represent the difference interquartile range and dots show the mean difference. No significant differences were found for any measurements using linear mixed models.

**Table S4. OCT volumes significance table (scan 1 — scan 2).** Significance values for OCT volumes measures comparisons between repeated and original scans.

scan 1 — scan 2

| | | p-value | $\beta_0$ | $\beta_1$ | 2.5 % lower CI bound | 97.5 % upper CI bound |
|---|---|---|---|---|---|---|
| CV | global | 8.0369E-01 | 8.7883 | 0.9046 | -0.0985 | 0.0768 |
| | C | 6.5783E-01 | 0.2338 | 0.0430 | -0.0079 | 0.0050 |
| | SI | 4.3515E-01 | 0.4741 | 0.0703 | -0.0271 | 0.0116 |
| | NI | 9.2204E-02 | 0.4359 | 0.0844 | -0.0225 | 0.0002 |
| | II | 4.5899E-01 | 0.4453 | 0.0870 | -0.0092 | 0.0042 |
| | TI | 4.3950E-01 | 0.4534 | 0.0706 | -0.0164 | 0.0071 |
| | SO | 6.1303E-01 | 1.4703 | 0.1769 | -0.0132 | 0.0222 |
| | NO | 8.1137E-01 | 1.1462 | 0.1964 | -0.0392 | 0.0309 |
| | IO | 5.0251E-01 | 1.3187 | 0.1740 | -0.0151 | 0.0307 |
| | TO | 2.4469E-01 | 1.3414 | 0.1792 | -0.0332 | 0.0077 |
| LV | global | 4.6223E-01 | 3.8096 | 0.5421 | -0.0766 | 0.0348 |
| | C | 3.0986E-01 | 0.1110 | 0.0269 | -0.0078 | 0.0024 |
| | SI | 5.0852E-01 | 0.2307 | 0.0491 | -0.0211 | 0.0105 |
| | NI | 6.8924E-02 | 0.2043 | 0.0497 | -0.0181 | -0.0008 |
| | II | 5.3679E-01 | 0.2091 | 0.0534 | -0.0078 | 0.0041 |
| | TI | 5.0001E-01 | 0.2056 | 0.0435 | -0.0102 | 0.0050 |
| | SO | 5.4766E-01 | 0.6306 | 0.1151 | -0.0114 | 0.0214 |
| | NO | 6.9432E-01 | 0.4848 | 0.1132 | -0.0206 | 0.0138 |
| | IO | 2.7959E-01 | 0.5663 | 0.0940 | -0.0058 | 0.0215 |
| | TO | 3.3869E-01 | 0.5498 | 0.0842 | -0.0255 | 0.0085 |
| CT | global | 6.2949E-01 | 258.0147 | 36.5353 | -3.1089 | 1.8955 |
| | C | 4.0853E-01 | 297.5638 | 54.2303 | -15.0940 | 6.0786 |
| | SI | 5.3990E-01 | 301.4390 | 45.1992 | -18.2263 | 9.5895 |
| | NI | 1.6564E-01 | 276.7596 | 53.9831 | -15.1766 | 2.0097 |
| | II | 5.9570E-01 | 283.1461 | 55.5973 | -5.6594 | 3.2714 |
| | TI | 5.4838E-01 | 287.8310 | 45.1268 | -9.9663 | 5.3221 |
| | SO | 4.0494E-01 | 282.3029 | 35.3242 | -2.4303 | 6.0871 |
| | NO | 9.8604E-01 | 220.5788 | 38.7028 | -7.2111 | 7.0869 |
| | IO | 3.2690E-01 | 254.2534 | 35.6720 | -2.2764 | 7.0722 |
| | TO | 4.7373E-01 | 258.3787 | 37.5059 | -5.6080 | 2.6049 |
| LT | global | 3.9451E-01 | 112.1764 | 19.8597 | -2.2869 | 0.8900 |
| | C | 2.7234E-01 | 141.2867 | 34.0209 | -12.6698 | 3.3447 |
| | SI | 5.5872E-01 | 146.7434 | 31.4532 | -13.9631 | 7.5906 |
| | NI | 9.5439E-02 | 129.7710 | 31.7063 | -11.8688 | 0.1879 |
| | II | 5.7232E-01 | 133.0419 | 34.0532 | -4.9039 | 2.7279 |
| | TI | 5.6250E-01 | 130.5610 | 27.6817 | -6.4887 | 3.5502 |
| | SO | 4.5354E-01 | 121.1375 | 22.5219 | -2.1686 | 4.8676 |
| | NO | 8.2072E-01 | 93.3072 | 21.9945 | -3.8442 | 3.0649 |
| | IO | 2.1446E-01 | 109.2763 | 18.9633 | -0.9206 | 4.6791 |
| | TO | 4.7296E-01 | 105.9392 | 17.4588 | -4.5825 | 2.1253 |
| CVI | global | 2.2243E-01 | 0.4290 | 0.0189 | -0.0042 | 0.0009 |
| | C | 1.4507E-01 | 0.4599 | 0.0390 | -0.0189 | 0.0019 |
| | SI | 6.7827E-01 | 0.4734 | 0.0382 | -0.0140 | 0.0092 |
| | NI | 8.0554E-02 | 0.4535 | 0.0312 | -0.0141 | -0.0002 |
| | II | 6.1587E-01 | 0.4539 | 0.0383 | -0.0094 | 0.0056 |
| | TI | 7.6407E-01 | 0.4439 | 0.0325 | -0.0088 | 0.0065 |
| | SO | 4.4242E-01 | 0.4213 | 0.0272 | -0.0036 | 0.0083 |
| | NO | 9.5496E-01 | 0.4129 | 0.0269 | -0.0084 | 0.0080 |
| | IO | 9.4881E-02 | 0.4244 | 0.0164 | -0.0001 | 0.0073 |
| | TO | 4.0713E-01 | 0.4078 | 0.0090 | -0.0102 | 0.0041 |

**Table S5. OCT volumes repeatability metrics table (scan 1 — scan 2).** Intraclass correlation coefficients (ICC) and standard error of measurement (SEM) for OCT volumes measures comparisons between repeated and original scans.

scan 1 — scan 2

| | | ICC | SEM |
|---|---|---|---|
| CV | global | 0.9980 | 0.0628 |
| | C | 0.9919 | 0.0063 |
| | SI | 0.9700 | 0.0198 |
| | NI | 0.9908 | 0.0142 |
| | II | 0.9998 | 0.0022 |
| | TI | 0.9902 | 0.0093 |
| | SO | 0.9950 | 0.0178 |
| | NO | 0.9959 | 0.0257 |
| | IO | 0.9975 | 0.0148 |
| | TO | 0.9924 | 0.0214 |
| LV | global | 0.9982 | 0.0388 |
| | C | 0.9826 | 0.0060 |
| | SI | 0.9647 | 0.0161 |
| | NI | 0.9878 | 0.0106 |
| | II | 0.9996 | 0.0020 |
| | TI | 0.9958 | 0.0038 |
| | SO | 0.9923 | 0.0165 |
| | NO | 0.9985 | 0.0093 |
| | IO | 0.9972 | 0.0093 |
| | TO | 0.9821 | 0.0163 |
| CT | global | 0.9990 | 1.7593 |
| | C | 0.9823 | 11.7366 |
| | SI | 0.9641 | 13.9855 |
| | NI | 0.9894 | 9.8002 |
| | II | 0.9995 | 2.2111 |
| | TI | 0.9909 | 5.7578 |
| | SO | 0.9920 | 4.5363 |
| | NO | 0.9947 | 5.6881 |
| | IO | 0.9959 | 3.8499 |
| | TO | 0.9947 | 3.7383 |
| LT | global | 0.9992 | 0.9375 |
| | C | 0.9719 | 9.6378 |
| | SI | 0.9602 | 10.9974 |
| | NI | 0.9869 | 7.0160 |
| | II | 0.9994 | 1.5163 |
| | TI | 0.9951 | 2.6500 |
| | SO | 0.9900 | 3.6777 |
| | NO | 0.9981 | 2.0307 |
| | IO | 0.9957 | 2.2540 |
| | TO | 0.9851 | 3.0525 |
| CVI | global | 0.9984 | 0.0014 |
| | C | 0.9683 | 0.0113 |
| | SI | 0.9716 | 0.0113 |
| | NI | 0.9900 | 0.0065 |
| | II | 0.9971 | 0.0037 |
| | TI | 0.9911 | 0.0045 |
| | SO | 0.9833 | 0.0062 |
| | NO | 0.9967 | 0.0033 |
| | IO | 0.9915 | 0.0031 |
| | TO | 0.8585 | 0.0060 |

#### L. Additional temporal changes

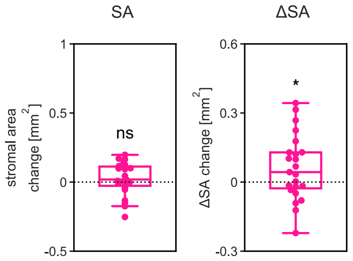


**Figure S16. Temporal choroid stromal pulsatile changes.** Difference box plots between post- and preflight SA and ΔSA measurements obtained over the selected segments for all astronaut eyes. ns nonsignificant, *P $<$ 0.05, linear mixed model.

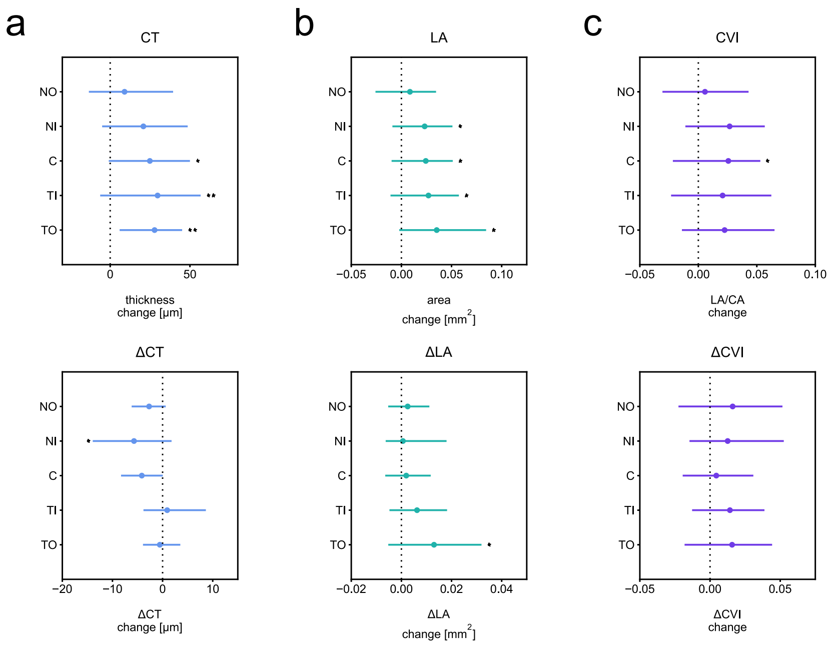


**Figure S17. Pre-spaceflight and post-spaceflight temporal quantification difference plots.** (**a**) Difference plot between post- and preflight CT and ΔCT measurements obtained over the selected segments for all astronaut eyes. Cross-sectional ETDRS subfield differences are shown. Difference plot lines represent the difference interquartile range and dots show the mean difference. (**b**) Difference plot between post- and preflight LA and ΔLA measurements. (**c**) Difference plot between post- and preflight CVI and ΔCVI measurements. *P $<$ 0.05, **P $<$ 0.01, linear mixed model.

#### M. Pre-spaceflight and post-spaceflight sample timeseries and transforms

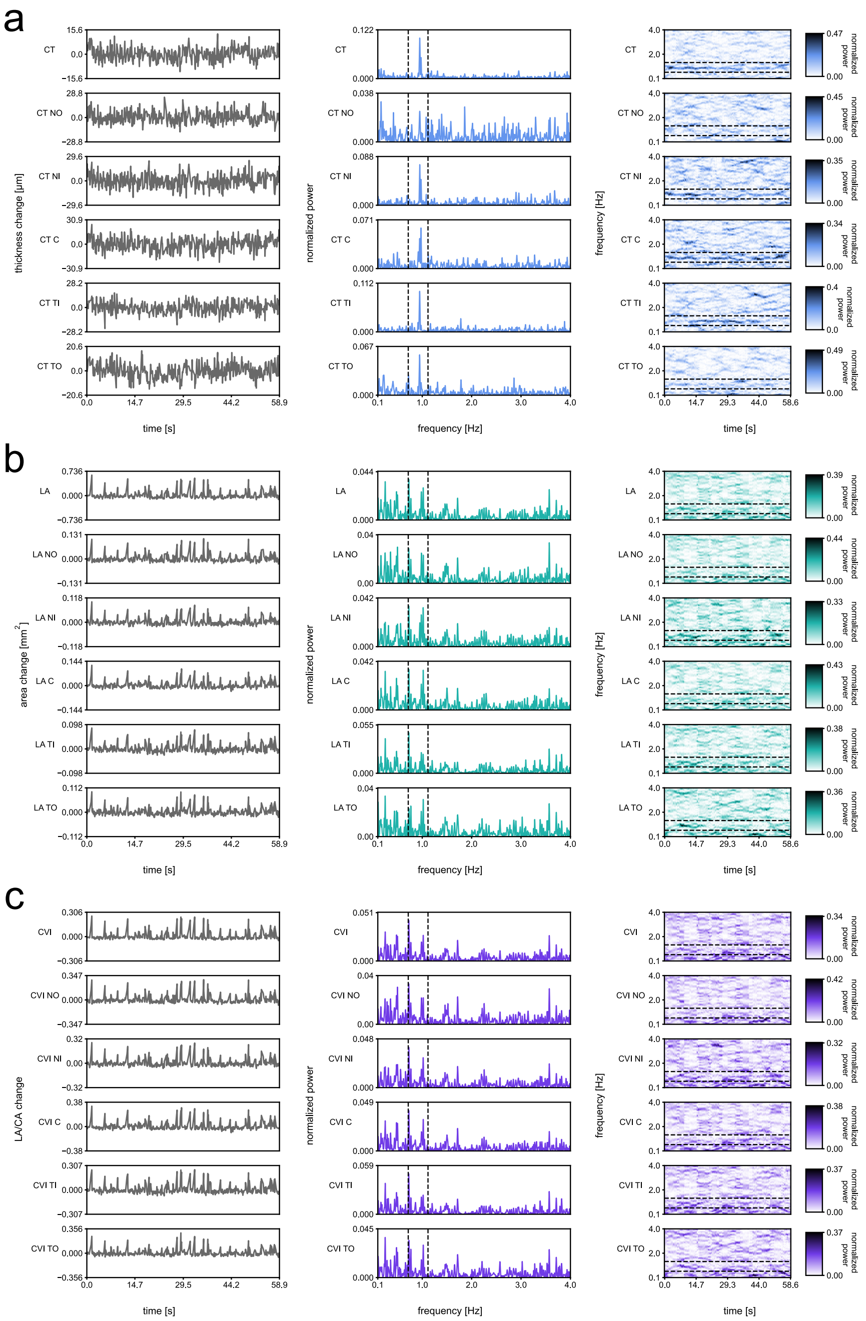


**Figure S18. Pre-spaceflight sample timeseries and transforms.** (**a**) Pre-spaceflight CT selected segment timeseries, frequency and joint time-frequency transforms for one subject's eye acquisition. First column features selected segment timeseries, second column features Lomb–Scargle periodograms and third column features Lomb–Scargle spectrograms. Dashed lines on the Lomb–Scargle periodogram and Lomb–Scargle spectrogram show values ± 20 % and ± 40 % of the oximeter measured heart rate value, respectively. (**b**) Pre-spaceflight LA selected segment timeseries, frequency and joint time-frequency transforms for the same subject's eye acquisition. (**c**) Pre-spaceflight CVI selected segment timeseries, frequency and joint time-frequency transforms for the same subject's eye acquisition.

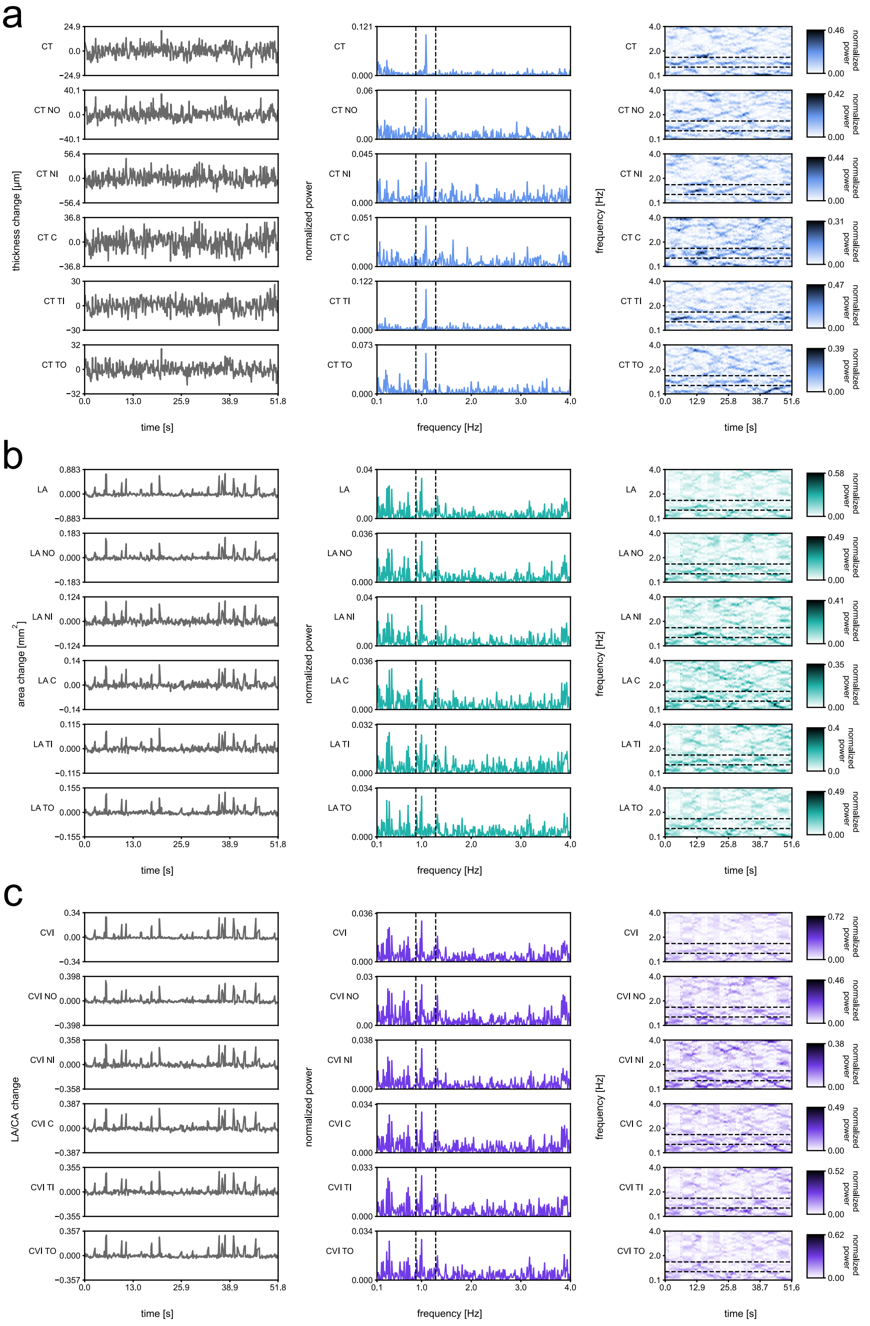


**Figure S19. Post-spaceflight sample timeseries and transforms.** (**a**) Post-spaceflight CT selected segment timeseries, frequency and joint time-frequency transforms for one subject's eye acquisition. First column features selected segment timeseries, second column features Lomb–Scargle periodograms and third column features Lomb–Scargle spectrograms. Dashed lines on the Lomb–Scargle periodogram and Lomb–Scargle spectrogram show values $\pm$ 20 % and $\pm$ 40 % of the oximeter measured heart rate value, respectively. (**b**) Post-spaceflight LA selected segment timeseries, frequency and joint time-frequency transforms for the same subject's eye acquisition. (**c**) Post-spaceflight CVI selected segment timeseries, frequency and joint time-frequency transforms for the same subject's eye acquisition.

#### N. Additional spatial changes

One or both eyes from all 6 subjects were represented in at least one preflight timepoint ($n_{eye}$ = 10, $n_{subject}$ = 5 for launch - 21 to 18 months; $n_{eye}$ = 11, $n_{subject}$ = 6 for launch - 9 to 6 months). Not all eyes from all six subjects were available for individual inflight and postflight timepoints. When obtaining the significance of differences between individual inflight and postflight timepoint quantifications and their preflight baseline, if an eye was represented in both preflight timepoints, the two preflight quantification values were averaged. Quantification values corresponding to the available eyes for individual inflight and postflight timepoints were then compared with the preflight baseline ($n_{eye}$ = 11, $n_{subject}$ = 6 for launch + 30 days; $n_{eye}$ = 10, $n_{subject}$ = 5 for launch + 90 days; $n_{eye}$ = 6, $n_{subject}$ = 3 for return - 30 days; $n_{eye}$ = 10, $n_{subject}$ = 5 for return + 1 to 3 days).

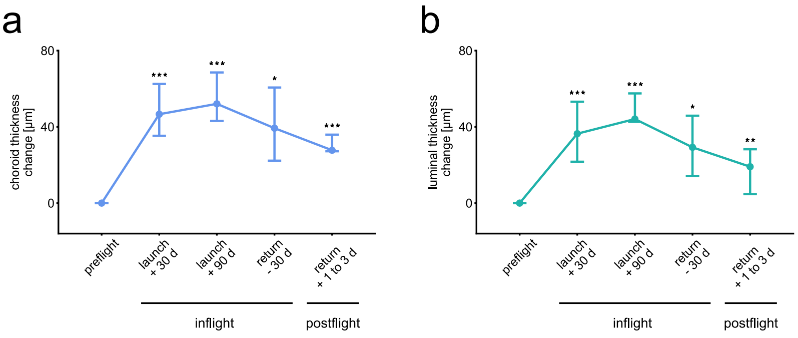


**Figure S20. Spatial choroid thickness changes.** (**a**) Line chart of change in CT over preflight, inflight and postflight timepoints for all astronaut eyes. Error bars show the difference interquartile range and dots show the mean difference. (**b**) Line chart of change in LT over preflight, inflight and postflight timepoints for the same eyes. (d days). *P $<$ 0.05, **P $<$ 0.01, ***P $<$ 0.001, linear mixed model.

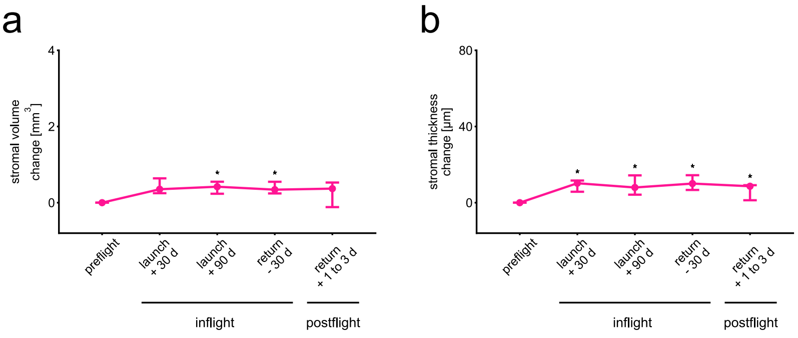


**Figure S21. Spatial choroid stromal volume and thickness changes.** (**a**) Line chart of change in stromal volume and (**b**) thickness over preflight, inflight and postflight timepoints for all astronaut eyes. Error bars show the difference interquartile range and dots show the mean difference. (d days). *P $<$ 0.05, linear mixed model.

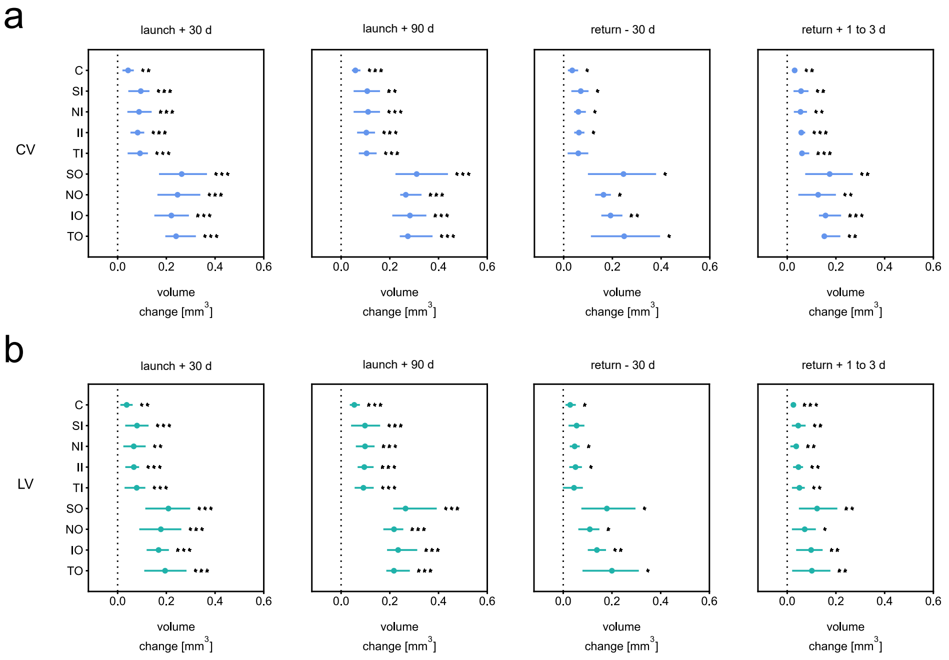


**Figure S22. Spatial quantification choroid volume difference plots.** (**a**) Difference plots between several timepoints and preflight CV measurements over the specified macular regions for all astronaut eyes. ETDRS subfield differences are shown. Difference plot lines represent the difference interquartile range and dots show the mean difference. (**b**) Difference plot between several timepoints and preflight LV measurements. (d days). *$P < 0.05$, **$P < 0.01$, ***$P < 0.001$, linear mixed model.

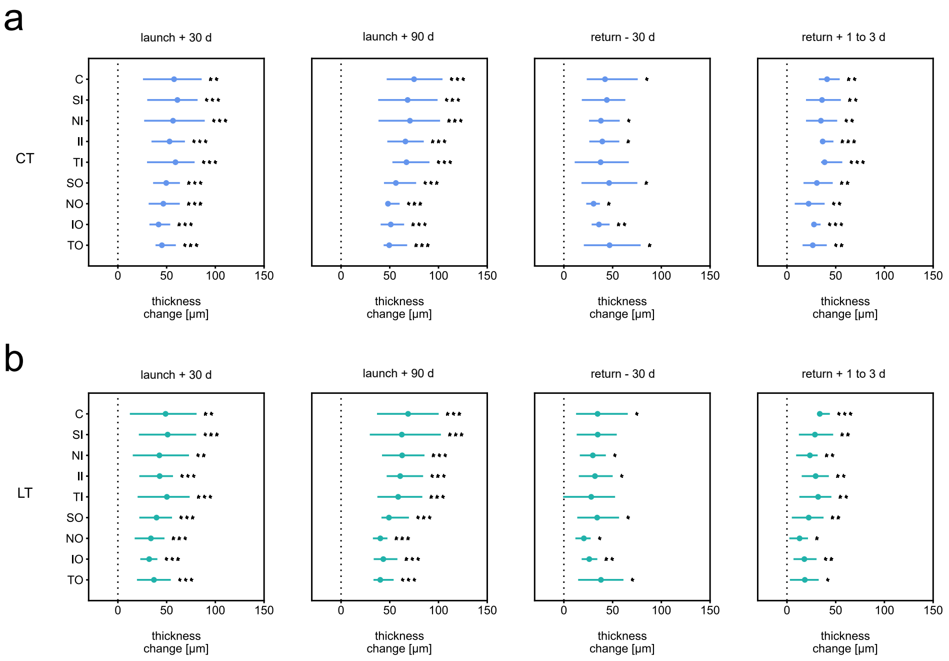


**Figure S23. Spatial quantification choroid thickness difference plots.** (**a**) Difference plots between several timepoints and preflight CT measurements averaged over the specified macular regions for all astronaut eyes. ETDRS subfield differences are shown. Difference plot lines represent the difference interquartile range and dots show the mean difference. (**b**) Difference plots between several timepoints and preflight LT measurements. (d days). *$P < 0.05$, **$P < 0.01$, ***$P < 0.001$, linear mixed model.

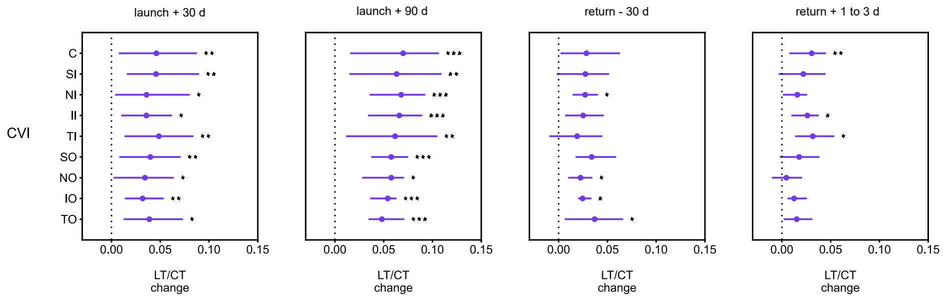


**Figure S24. Spatial quantification CVI difference plots.** Difference plots between several timepoints and preflight CVI measurements averaged over the specified macular regions for all astronaut eyes. ETDRS subfield differences are shown. Difference plot lines represent the difference interquartile range and dots show the mean difference. (d days). *$P < 0.05$, **$P < 0.01$, ***$P < 0.001$, linear mixed model.

#### O. Pre-spaceflight spatial changes

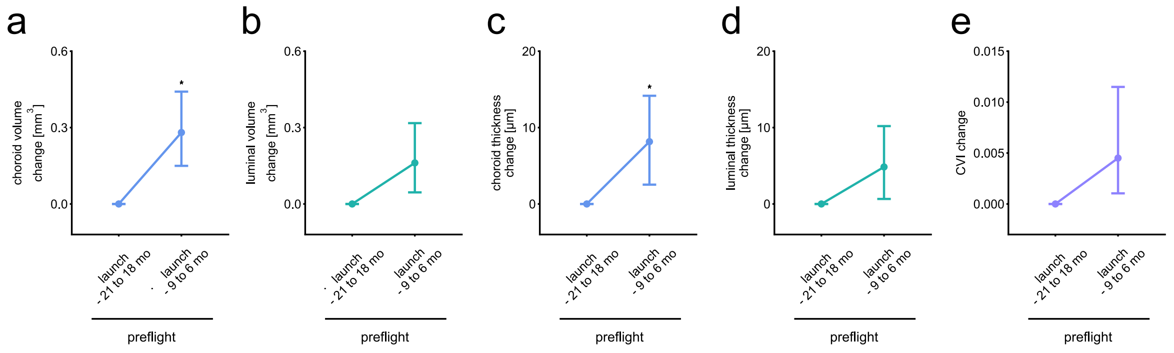


**Figure S25. Pre-spaceflight spatial OCT choroid changes.** (**a**) Line chart of change in CV over preflight timepoints for all OCT volumes astronaut eyes. Error bars show the difference interquartile range and dots show the mean difference. (**b**) Line chart of change in LV. (**c**) Line chart of change in CT. (**d**) Line chart of change in LT. (**e**) Line chart of change in CVI. (d days). *P $<$ 0.05, linear mixed model.

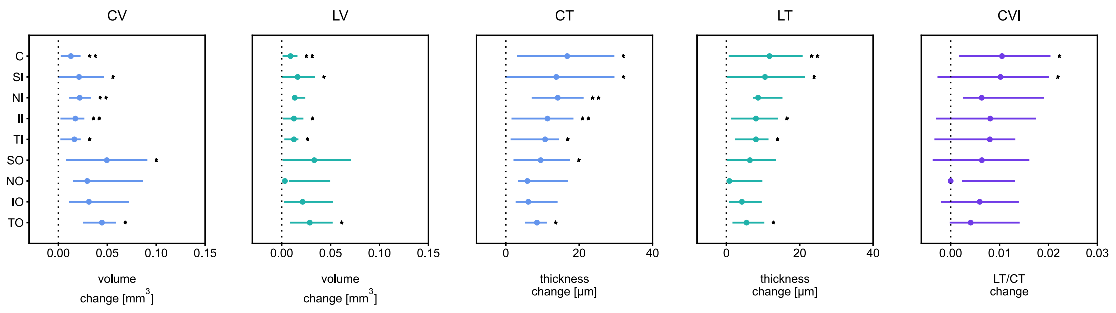


**Figure S26. Pre-spaceflight spatial quantification difference plots.** Difference plots between preflight timepoints CV and LV measurements over the specified macular regions, and CT, LT and CVI measurements averaged over the specified macular regions for all astronaut eyes. ETDRS subfield differences are shown. Difference plot lines represent the difference interquartile range and dots show the mean difference. *P $<$ 0.05, **P $<$ 0.01, linear mixed model.

#### P. Quantification tables

**Table S6. OCT videos quantification table.** Quantification values for OCT videos at the preflight timepoint (average ± standard deviation).

| | CT [μm] | ΔCT [μm] | LA [mm$^2$] | ΔLA [mm$^2$] | CVI | ΔCVI |
|---|---|---|---|---|---|---|
| global | 289.8±89.4 | 15.146±6.213 | 1.569±0.622 | 0.2303±0.1006 | 0.5985±0.0769 | 0.0901±0.0406 |
| NO | 270.4±102.9 | 19.908±8.831 | 0.245±0.129 | 0.0451±0.0267 | 0.5786±0.0909 | 0.096±0.0454 |
| NI | 339.8±119.8 | 29.527±15.978 | 0.221±0.103 | 0.0419±0.0175 | 0.6229±0.093 | 0.1022±0.0418 |
| C | 360.8±123.7 | 31.641±16.974 | 0.238±0.108 | 0.0443±0.017 | 0.6292±0.1031 | 0.103±0.0399 |
| TI | 345±111.8 | 27.619±16.66 | 0.227±0.098 | 0.0401±0.0167 | 0.634±0.0953 | 0.0961±0.0368 |
| TO | 302.7±92.7 | 23.517±14.239 | 0.275±0.11 | 0.0518±0.0227 | 0.5907±0.0807 | 0.1025±0.0451 |

**Table S7. OCT volumes quantification table.** Quantification values for OCT volumes at the preflight timepoint (average ± standard deviation).

| | CV [mm$^3$] | LV [mm$^3$] | CT [μm] | LT [μm] | CVI |
|---|---|---|---|---|---|
| global | 10.7893±2.8846 | 5.2799±2.1061 | 309.2±75.6 | 150.7±54.9 | 0.4743±0.0623 |
| C | 0.2792±0.0739 | 0.1495±0.0619 | 355.5±94.8 | 190.4±79.4 | 0.5135±0.0936 |
| SI | 0.555±0.135 | 0.2997±0.1139 | 351.5±84.2 | 189.7±71.3 | 0.5218±0.084 |
| NI | 0.5275±0.1521 | 0.2763±0.1172 | 333.6±95 | 174.7±73.4 | 0.5024±0.0841 |
| II | 0.5447±0.1528 | 0.2901±0.1202 | 344.9±95.5 | 183.6±75.3 | 0.511±0.0878 |
| TI | 0.5401±0.1422 | 0.2798±0.1189 | 341.6±88.5 | 176.8±74.3 | 0.4976±0.0884 |
| SO | 1.7037±0.3809 | 0.8224±0.2945 | 326.1±70.8 | 157.2±54.8 | 0.4699±0.0653 |
| NO | 1.4174±0.4569 | 0.6724±0.3106 | 271.2±84.7 | 128.5±57.8 | 0.4561±0.063 |
| IO | 1.6603±0.462 | 0.8204±0.338 | 318.3±85.6 | 157±62.6 | 0.4777±0.0668 |
| TO | 1.6327±0.423 | 0.7625±0.301 | 312.8±78.7 | 145.8±55.8 | 0.4531±0.063 |

#### Q. Significance tables

**Table S8. OCT videos significance table (preflight — postflight).** Significance values for OCT videos measures comparisons between pre- and postflight.

preflight — postflight

| | | p-value | $\beta_0$ | $\beta_1$ | 2.5 % lower CI bound | 97.5 % upper CI bound |
|---|---|---|---|---|---|---|
| CT | global | 2.0686E-02 | 290.7543 | 24.2108 | 3.0407 | 27.7030 |
| | NO | 1.7714E-01 | 272.9097 | 27.7098 | -3.9107 | 21.9229 |
| | NI | 5.1475E-02 | 343.8202 | 32.8081 | 0.6032 | 41.0096 |
| | C | 2.1198E-02 | 364.2329 | 33.3269 | 4.8271 | 44.8908 |
| | TI | 4.9417E-03 | 346.9840 | 30.8648 | 10.6939 | 48.7443 |
| | TO | 4.3978E-03 | 301.6968 | 25.2965 | 10.2888 | 45.3483 |
| ΔCT | global | 2.9872E-01 | 15.3387 | 1.4220 | -3.0668 | 0.9169 |
| | NO | 5.9779E-02 | 20.0871 | 2.0221 | -5.3857 | 0.0024 |
| | NI | 2.2842E-02 | 30.0959 | 3.4738 | -10.3630 | -1.0438 |
| | C | 9.7902E-02 | 32.7005 | 4.1550 | -8.9450 | 0.6504 |
| | TI | 7.8396E-01 | 28.1478 | 3.6750 | -5.5823 | 7.4101 |
| | TO | 8.3352E-01 | 23.5325 | 3.2809 | -5.7986 | 4.6836 |
| LA | global | 2.9313E-02 | 1.5784 | 0.1668 | 0.0176 | 0.2296 |
| | NO | 3.5620E-01 | 0.2484 | 0.0333 | -0.0095 | 0.0266 |
| | NI | 3.1106E-02 | 0.2239 | 0.0282 | 0.0031 | 0.0433 |
| | C | 2.4453E-02 | 0.2416 | 0.0300 | 0.0042 | 0.0444 |
| | TI | 1.9946E-02 | 0.2291 | 0.0275 | 0.0055 | 0.0485 |
| | TO | 2.4559E-02 | 0.2734 | 0.0295 | 0.0061 | 0.0643 |
| ΔLA | global | 2.8249E-02 | 0.2302 | 0.0299 | 0.0093 | 0.1065 |
| | NO | 6.3332E-01 | 0.0451 | 0.0069 | -0.0077 | 0.0127 |
| | NI | 8.5690E-01 | 0.0422 | 0.0047 | -0.0060 | 0.0072 |
| | C | 5.6716E-01 | 0.0450 | 0.0046 | -0.0047 | 0.0085 |
| | TI | 1.3476E-01 | 0.0403 | 0.0041 | -0.0017 | 0.0141 |
| | TO | 3.7293E-02 | 0.0518 | 0.0061 | 0.0014 | 0.0246 |
| CVI | global | 1.9770E-01 | 0.6004 | 0.0197 | -0.0073 | 0.0361 |
| | NO | 5.7479E-01 | 0.5814 | 0.0225 | -0.0143 | 0.0256 |
| | NI | 6.2884E-02 | 0.6256 | 0.0246 | -0.0006 | 0.0540 |
| | C | 3.9220E-02 | 0.6333 | 0.0280 | 0.0026 | 0.0487 |
| | TI | 2.0262E-01 | 0.6366 | 0.0256 | -0.0106 | 0.0521 |
| | TO | 1.8544E-01 | 0.5905 | 0.0181 | -0.0104 | 0.0553 |
| ΔCVI | global | 7.0513E-02 | 0.0895 | 0.0113 | -0.0009 | 0.0432 |
| | NO | 2.0425E-01 | 0.0953 | 0.0123 | -0.0082 | 0.0404 |
| | NI | 3.0919E-01 | 0.1012 | 0.0130 | -0.0114 | 0.0365 |
| | C | 6.8890E-01 | 0.1022 | 0.0113 | -0.0174 | 0.0263 |
| | TI | 2.2723E-01 | 0.0957 | 0.0106 | -0.0085 | 0.0368 |
| | TO | 1.9378E-01 | 0.1020 | 0.0125 | -0.0076 | 0.0390 |

**Table S9. OCT volumes significance table (preflight — launch + 30 days).** Significance values for OCT volumes measures comparisons between preflight and launch + 30 days.

preflight — L + 30 days

| | | p-value | $\beta_0$ | $\beta_1$ | 2.5 % lower CI bound | 97.5 % upper CI bound |
|---|---|---|---|---|---|---|
| CV | global | 1.3192E-04 | 10.8851 | 1.1881 | 1.0375 | 2.1098 |
| | C | 1.4391E-03 | 0.2814 | 0.0317 | 0.0231 | 0.0630 |
| | SI | 3.3652E-04 | 0.5580 | 0.0568 | 0.0586 | 0.1317 |
| | NI | 2.7109E-04 | 0.5368 | 0.0664 | 0.0551 | 0.1208 |
| | II | 2.4154E-05 | 0.5534 | 0.0657 | 0.0594 | 0.1051 |
| | TI | 2.2318E-04 | 0.5432 | 0.0609 | 0.0585 | 0.1255 |
| | SO | 4.1286E-05 | 1.7123 | 0.1654 | 0.1852 | 0.3408 |
| | NO | 2.3842E-05 | 1.4390 | 0.2024 | 0.1778 | 0.3143 |
| | IO | 4.2335E-06 | 1.6779 | 0.1908 | 0.1703 | 0.2708 |
| | TO | 5.4938E-05 | 1.6405 | 0.1877 | 0.1665 | 0.3134 |
| LV | global | 1.3279E-04 | 5.3635 | 0.9096 | 0.8046 | 1.6371 |
| | C | 2.5020E-03 | 0.1521 | 0.0262 | 0.0185 | 0.0559 |
| | SI | 7.2407E-04 | 0.3034 | 0.0482 | 0.0457 | 0.1135 |
| | NI | 1.7375E-03 | 0.2848 | 0.0505 | 0.0342 | 0.0982 |
| | II | 1.5383E-04 | 0.2976 | 0.0521 | 0.0435 | 0.0897 |
| | TI | 6.0565E-04 | 0.2831 | 0.0509 | 0.0458 | 0.1109 |
| | SO | 2.1064E-04 | 0.8338 | 0.1342 | 0.1332 | 0.2839 |
| | NO | 5.2764E-04 | 0.6892 | 0.1470 | 0.1051 | 0.2499 |
| | IO | 1.3782E-05 | 0.8342 | 0.1458 | 0.1244 | 0.2120 |
| | TO | 4.8753E-04 | 0.7694 | 0.1394 | 0.1163 | 0.2736 |
| CT | global | 1.0347E-05 | 311.7816 | 33.6692 | 34.8815 | 58.3614 |
| | C | 1.1073E-03 | 358.6174 | 40.7529 | 32.3037 | 83.0070 |
| | SI | 3.0831E-04 | 353.2410 | 35.8903 | 37.8943 | 84.2895 |
| | NI | 2.1618E-04 | 339.3825 | 41.9562 | 36.0582 | 77.1018 |
| | II | 1.5696E-05 | 350.3320 | 41.5318 | 39.0130 | 67.0166 |
| | TI | 1.9366E-04 | 343.4600 | 38.4391 | 37.9693 | 80.2390 |
| | SO | 3.1986E-05 | 327.6975 | 31.5182 | 35.3710 | 63.8375 |
| | NO | 1.7052E-05 | 275.3513 | 38.1495 | 34.1975 | 59.0659 |
| | IO | 6.6267E-07 | 321.5776 | 35.9711 | 34.0039 | 49.5397 |
| | TO | 5.4825E-05 | 314.1396 | 35.4932 | 31.3487 | 58.9851 |
| LT | global | 1.1454E-04 | 153.0361 | 25.2793 | 24.2187 | 48.5904 |
| | C | 2.1433E-03 | 193.9014 | 33.6313 | 24.8728 | 72.9841 |
| | SI | 6.6033E-04 | 191.9606 | 30.4441 | 29.5898 | 72.5440 |
| | NI | 1.5191E-03 | 179.9304 | 31.9163 | 22.4150 | 62.7496 |
| | II | 1.2946E-04 | 188.2563 | 32.9343 | 28.2965 | 57.4350 |
| | TI | 5.4520E-04 | 178.8583 | 32.1009 | 29.6846 | 70.8863 |
| | SO | 1.9014E-04 | 159.3992 | 25.3795 | 25.5003 | 53.7856 |
| | NO | 5.4427E-04 | 131.6881 | 27.7271 | 20.0036 | 47.7593 |
| | IO | 1.1092E-05 | 159.6220 | 27.3839 | 23.9849 | 40.3019 |
| | TO | 5.5395E-04 | 147.0821 | 26.1806 | 21.8193 | 52.2171 |
| CVI | global | 6.2860E-03 | 0.4776 | 0.0282 | 0.0147 | 0.0577 |
| | C | 7.7744E-03 | 0.5179 | 0.0392 | 0.0181 | 0.0740 |
| | SI | 6.0967E-03 | 0.5248 | 0.0353 | 0.0187 | 0.0727 |
| | NI | 2.9470E-02 | 0.5081 | 0.0348 | 0.0072 | 0.0647 |
| | II | 1.0049E-02 | 0.5168 | 0.0378 | 0.0127 | 0.0588 |
| | TI | 8.4536E-03 | 0.5008 | 0.0375 | 0.0182 | 0.0790 |
| | SO | 5.7161E-03 | 0.4734 | 0.0295 | 0.0166 | 0.0631 |
| | NO | 1.3139E-02 | 0.4593 | 0.0299 | 0.0110 | 0.0577 |
| | IO | 3.1398E-03 | 0.4810 | 0.0291 | 0.0151 | 0.0489 |
| | TO | 1.0153E-02 | 0.4558 | 0.0292 | 0.0137 | 0.0640 |

**Table S10. OCT volumes significance table (preflight — launch + 90 days).** Significance values for OCT volumes measures comparisons between preflight and launch + 90 days.

preflight — L + 90 days

| | | p-value | $\beta_0$ | $\beta_1$ | 2.5 % lower CI bound | 97.5 % upper CI bound |
|---|---|---|---|---|---|---|
| CV | global | 1.9766E-04 | 9.8790 | 1.1207 | 1.3814 | 2.7787 |
| | C | 2.7162E-04 | 0.2651 | 0.0389 | 0.0385 | 0.0790 |
| | SI | 1.0846E-03 | 0.5239 | 0.0634 | 0.0615 | 0.1533 |
| | NI | 3.5153E-04 | 0.4900 | 0.0716 | 0.0699 | 0.1516 |
| | II | 2.3588E-04 | 0.5119 | 0.0747 | 0.0679 | 0.1395 |
| | TI | 7.8115E-04 | 0.5127 | 0.0717 | 0.0622 | 0.1476 |
| | SO | 4.6934E-04 | 1.5854 | 0.1553 | 0.1909 | 0.4295 |
| | NO | 3.6427E-05 | 1.2598 | 0.1569 | 0.1930 | 0.3381 |
| | IO | 6.6473E-05 | 1.5154 | 0.1786 | 0.1992 | 0.3661 |
| | TO | 5.0227E-04 | 1.5265 | 0.1957 | 0.1677 | 0.3806 |
| LV | global | 7.8630E-05 | 4.6209 | 0.8474 | 1.1673 | 2.1540 |
| | C | 1.5331E-04 | 0.1376 | 0.0313 | 0.0366 | 0.0717 |
| | SI | 7.1859E-04 | 0.2749 | 0.0530 | 0.0585 | 0.1375 |
| | NI | 3.5394E-04 | 0.2460 | 0.0541 | 0.0626 | 0.1340 |
| | II | 1.8608E-04 | 0.2653 | 0.0593 | 0.0635 | 0.1270 |
| | TI | 4.0332E-04 | 0.2598 | 0.0582 | 0.0577 | 0.1257 |
| | SO | 3.8510E-04 | 0.7293 | 0.1187 | 0.1688 | 0.3604 |
| | NO | 6.9655E-04 | 0.5596 | 0.1126 | 0.1288 | 0.3057 |
| | IO | 8.8962E-05 | 0.7127 | 0.1328 | 0.1624 | 0.3059 |
| | TO | 2.4355E-04 | 0.6862 | 0.1360 | 0.1419 | 0.2918 |
| CT | global | 1.4107E-04 | 286.3574 | 32.1486 | 35.1002 | 69.0443 |
| | C | 3.2482E-04 | 337.3298 | 49.3794 | 47.9769 | 101.8424 |
| | SI | 9.5487E-04 | 332.3974 | 40.0250 | 39.5796 | 97.0588 |
| | NI | 2.6447E-04 | 310.3498 | 45.1673 | 45.6136 | 95.8011 |
| | II | 1.6465E-04 | 324.6697 | 47.2299 | 44.1347 | 87.9637 |
| | TI | 6.4491E-04 | 324.7206 | 45.2114 | 40.4661 | 93.7564 |
| | SO | 3.5072E-04 | 304.8400 | 30.1070 | 35.4835 | 77.0003 |
| | NO | 2.5145E-05 | 242.4246 | 29.8249 | 35.6027 | 60.7215 |
| | IO | 5.1029E-05 | 292.0515 | 34.0215 | 36.4641 | 65.5753 |
| | TO | 4.9857E-04 | 293.8356 | 37.4624 | 30.2709 | 68.6407 |
| LT | global | 8.2857E-05 | 133.5264 | 22.8922 | 30.9053 | 57.2827 |
| | C | 2.0754E-04 | 175.2539 | 39.6568 | 45.5398 | 92.0507 |
| | SI | 6.2916E-04 | 174.3436 | 33.4904 | 37.7245 | 86.9687 |
| | NI | 3.0073E-04 | 155.7256 | 34.1562 | 40.4037 | 84.9933 |
| | II | 1.5691E-04 | 168.1382 | 37.4855 | 40.9492 | 80.4717 |
| | TI | 3.3072E-04 | 164.3820 | 36.7267 | 37.4855 | 79.7370 |
| | SO | 3.5957E-04 | 140.1701 | 22.6485 | 31.2276 | 67.0767 |
| | NO | 8.3113E-04 | 107.6082 | 21.2084 | 23.4719 | 57.1705 |
| | IO | 9.9470E-05 | 137.1957 | 24.9577 | 29.8805 | 56.8846 |
| | TO | 2.8855E-04 | 131.8969 | 25.6764 | 26.0203 | 54.4970 |
| CVI | global | 1.5056E-04 | 0.4577 | 0.0287 | 0.0362 | 0.0709 |
| | C | 6.1028E-04 | 0.4972 | 0.0442 | 0.0424 | 0.0974 |
| | SI | 1.6053E-03 | 0.5077 | 0.0402 | 0.0345 | 0.0920 |
| | NI | 3.6447E-04 | 0.4832 | 0.0389 | 0.0431 | 0.0926 |
| | II | 2.1042E-04 | 0.4966 | 0.0426 | 0.0436 | 0.0883 |
| | TI | 1.3189E-03 | 0.4858 | 0.0411 | 0.0345 | 0.0888 |
| | SO | 4.0053E-04 | 0.4526 | 0.0298 | 0.0363 | 0.0787 |
| | NO | 1.7561E-02 | 0.4353 | 0.0312 | 0.0172 | 0.0981 |
| | IO | 7.9094E-05 | 0.4594 | 0.0291 | 0.0382 | 0.0700 |
| | TO | 3.7087E-04 | 0.4388 | 0.0281 | 0.0305 | 0.0657 |

**Table S11. OCT volumes significance table (preflight — return - 30 days).** Significance values for OCT volumes measures comparisons between preflight and return - 30 days.

preflight — R - 30 days

| | | p-value | $\beta_0$ | $\beta_1$ | 2.5 % lower CI bound | 97.5 % upper CI bound |
|---|---|---|---|---|---|---|
| CV | global | 2.0039E-02 | 9.9125 | 1.9894 | 0.4986 | 2.1939 |
| | C | 4.4913E-02 | 0.2620 | 0.0673 | 0.0080 | 0.0623 |
| | SI | 4.9036E-02 | 0.5128 | 0.1095 | 0.0127 | 0.1277 |
| | NI | 1.5815E-02 | 0.4753 | 0.1221 | 0.0247 | 0.0961 |
| | II | 1.4523E-02 | 0.5052 | 0.1317 | 0.0265 | 0.0989 |
| | TI | 6.7044E-02 | 0.5243 | 0.1242 | 0.0055 | 0.1145 |
| | SO | 2.8647E-02 | 1.4825 | 0.2283 | 0.0747 | 0.4165 |
| | NO | 1.4710E-02 | 1.1856 | 0.2495 | 0.0687 | 0.2584 |
| | IO | 4.3376E-03 | 1.4849 | 0.3128 | 0.1099 | 0.2751 |
| | TO | 2.9134E-02 | 1.5716 | 0.3518 | 0.0746 | 0.4220 |
| LV | global | 2.0571E-02 | 4.7151 | 1.4585 | 0.3842 | 1.6247 |
| | C | 4.5194E-02 | 0.1436 | 0.0541 | 0.0063 | 0.0507 |
| | SI | 5.9586E-02 | 0.2767 | 0.0916 | 0.0071 | 0.1028 |
| | NI | 1.8047E-02 | 0.2473 | 0.0915 | 0.0182 | 0.0757 |
| | II | 2.2873E-02 | 0.2762 | 0.1042 | 0.0183 | 0.0824 |
| | TI | 9.4182E-02 | 0.2843 | 0.0996 | 0.0000 | 0.0886 |
| | SO | 2.7613E-02 | 0.6668 | 0.1711 | 0.0558 | 0.3026 |
| | NO | 2.4414E-02 | 0.5153 | 0.1594 | 0.0366 | 0.1818 |
| | IO | 7.2383E-03 | 0.7145 | 0.2302 | 0.0712 | 0.2052 |
| | TO | 2.0797E-02 | 0.7386 | 0.2434 | 0.0759 | 0.3236 |
| CT | global | 1.7683E-02 | 276.3073 | 55.4411 | 15.3736 | 63.2153 |
| | C | 4.7227E-02 | 334.4942 | 85.3299 | 9.0194 | 75.4269 |
| | SI | 5.6801E-02 | 324.6893 | 69.0474 | 6.2048 | 81.9342 |
| | NI | 1.9735E-02 | 300.3977 | 76.8174 | 14.1678 | 61.8119 |
| | II | 1.6326E-02 | 319.7910 | 83.1041 | 15.9524 | 62.9266 |
| | TI | 7.0735E-02 | 331.4047 | 78.1882 | 2.8004 | 72.5620 |
| | SO | 3.0065E-02 | 282.4549 | 43.9212 | 13.6643 | 79.1761 |
| | NO | 1.7141E-02 | 226.0052 | 46.7933 | 12.0714 | 48.8497 |
| | IO | 5.5473E-03 | 283.3441 | 59.0061 | 19.6144 | 52.3288 |
| | TO | 3.0416E-02 | 299.8680 | 67.4897 | 13.6681 | 80.0998 |
| LT | global | 1.9205E-02 | 130.9883 | 39.1342 | 11.5433 | 46.9716 |
| | C | 4.8955E-02 | 183.5044 | 68.6603 | 7.0035 | 62.1432 |
| | SI | 6.3593E-02 | 175.0173 | 57.6736 | 3.8152 | 65.5976 |
| | NI | 2.0714E-02 | 156.1698 | 57.5029 | 10.8345 | 48.5836 |
| | II | 2.3754E-02 | 174.6762 | 65.6571 | 11.3817 | 52.4018 |
| | TI | 9.4028E-02 | 179.5333 | 62.6525 | -0.0050 | 56.1268 |
| | SO | 2.9057E-02 | 126.8746 | 32.3239 | 10.2725 | 58.0064 |
| | NO | 2.5808E-02 | 98.0980 | 29.7059 | 6.6541 | 34.3557 |
| | IO | 8.2254E-03 | 136.1471 | 43.0710 | 13.0190 | 39.0874 |
| | TO | 2.1130E-02 | 140.7073 | 46.0091 | 14.3577 | 61.7287 |
| CVI | global | 3.5450E-02 | 0.4593 | 0.0480 | 0.0078 | 0.0484 |
| | C | 1.3121E-01 | 0.5116 | 0.0779 | -0.0041 | 0.0610 |
| | SI | 1.4377E-01 | 0.5115 | 0.0690 | -0.0061 | 0.0610 |
| | NI | 2.4908E-02 | 0.4897 | 0.0647 | 0.0090 | 0.0453 |
| | II | 7.7523E-02 | 0.5105 | 0.0767 | 0.0017 | 0.0485 |
| | TI | 2.6726E-01 | 0.5097 | 0.0720 | -0.0124 | 0.0503 |
| | SO | 5.4953E-02 | 0.4392 | 0.0446 | 0.0051 | 0.0625 |
| | NO | 2.6793E-02 | 0.4227 | 0.0374 | 0.0071 | 0.0378 |
| | IO | 1.2612E-02 | 0.4627 | 0.0501 | 0.0109 | 0.0382 |
| | TO | 3.0015E-02 | 0.4540 | 0.0505 | 0.0115 | 0.0622 |

**Table S12. OCT volumes significance table (preflight — return + 1 to 3 days).** Significance values for OCT volumes measures comparisons between preflight and return + 1 to 3 days.

preflight — R + 1 to 3 days

| | | p-value | $\beta_0$ | $\beta_1$ | 2.5 % lower CI bound | 97.5 % upper CI bound |
|---|---|---|---|---|---|---|
| CV | global | 5.2933E-03 | 10.3116 | 1.3790 | 0.4586 | 1.6356 |
| | C | 1.6555E-03 | 0.2632 | 0.0362 | 0.0164 | 0.0448 |
| | SI | 1.9867E-03 | 0.5305 | 0.0660 | 0.0297 | 0.0840 |
| | NI | 1.8794E-03 | 0.5019 | 0.0768 | 0.0289 | 0.0807 |
| | II | 5.8985E-04 | 0.5153 | 0.0768 | 0.0348 | 0.0807 |
| | TI | 8.0393E-04 | 0.5054 | 0.0659 | 0.0355 | 0.0860 |
| | SO | 4.1051E-03 | 1.6875 | 0.2026 | 0.0806 | 0.2680 |
| | NO | 5.9033E-03 | 1.3857 | 0.2433 | 0.0542 | 0.1998 |
| | IO | 2.9975E-04 | 1.5947 | 0.2331 | 0.1005 | 0.2142 |
| | TO | 1.9637E-03 | 1.5516 | 0.2086 | 0.0798 | 0.2250 |
| LV | global | 2.1935E-03 | 4.8983 | 1.0239 | 0.3493 | 1.0068 |
| | C | 8.0536E-04 | 0.1327 | 0.0272 | 0.0149 | 0.0360 |
| | SI | 4.5895E-03 | 0.2732 | 0.0525 | 0.0205 | 0.0701 |
| | NI | 4.2907E-03 | 0.2522 | 0.0565 | 0.0169 | 0.0570 |
| | II | 1.5106E-03 | 0.2598 | 0.0556 | 0.0251 | 0.0674 |
| | TI | 2.8027E-03 | 0.2444 | 0.0495 | 0.0249 | 0.0756 |
| | SO | 7.0623E-03 | 0.8006 | 0.1578 | 0.0501 | 0.1953 |
| | NO | 1.4801E-02 | 0.6528 | 0.1715 | 0.0228 | 0.1210 |
| | IO | 1.5295E-03 | 0.7642 | 0.1708 | 0.0532 | 0.1432 |
| | TO | 8.6340E-03 | 0.6943 | 0.1432 | 0.0390 | 0.1634 |
| CT | global | 6.8128E-04 | 301.2666 | 41.0884 | 16.4730 | 38.9702 |
| | C | 1.4721E-03 | 334.5682 | 46.9172 | 22.4319 | 59.8651 |
| | SI | 1.5951E-03 | 336.5775 | 41.5389 | 19.3931 | 52.5854 |
| | NI | 1.4829E-03 | 318.0031 | 48.3772 | 18.9635 | 50.7489 |
| | II | 5.1841E-04 | 326.8474 | 48.4393 | 22.3655 | 50.9727 |
| | TI | 6.9516E-04 | 320.3291 | 41.5328 | 22.9173 | 54.3712 |
| | SO | 3.3384E-03 | 324.3653 | 38.4591 | 14.7434 | 46.6038 |
| | NO | 6.8588E-03 | 266.3682 | 45.8945 | 9.1377 | 35.2986 |
| | IO | 1.3731E-04 | 307.3305 | 44.2884 | 18.7225 | 36.7303 |
| | TO | 3.3703E-03 | 298.8997 | 39.9986 | 12.7131 | 40.2774 |
| LT | global | 4.0773E-03 | 142.7867 | 29.5612 | 8.8328 | 29.3364 |
| | C | 7.5698E-04 | 168.6344 | 35.1553 | 19.8024 | 47.5610 |
| | SI | 4.1349E-03 | 173.2606 | 33.0865 | 13.2483 | 44.1615 |
| | NI | 3.6451E-03 | 159.6913 | 35.5791 | 11.1437 | 35.9820 |
| | II | 1.4983E-03 | 164.6874 | 35.0925 | 15.9851 | 42.8575 |
| | TI | 2.7169E-03 | 154.8211 | 31.2288 | 15.9321 | 48.0491 |
| | SO | 9.3863E-03 | 153.7348 | 29.8599 | 8.3932 | 36.2683 |
| | NO | 2.4430E-02 | 125.2752 | 32.3683 | 3.1041 | 22.8349 |
| | IO | 4.0938E-03 | 147.0837 | 32.3211 | 8.2577 | 27.4549 |
| | TO | 1.5623E-02 | 133.5915 | 27.1990 | 5.6733 | 30.9943 |
| CVI | global | 1.4029E-01 | 0.4601 | 0.0306 | -0.0039 | 0.0327 |
| | C | 5.4877E-03 | 0.4861 | 0.0385 | 0.0133 | 0.0479 |
| | SI | 5.9404E-02 | 0.4986 | 0.0368 | 0.0011 | 0.0429 |
| | NI | 1.0217E-01 | 0.4816 | 0.0372 | -0.0020 | 0.0337 |
| | II | 2.9501E-02 | 0.4855 | 0.0365 | 0.0054 | 0.0469 |
| | TI | 2.0312E-02 | 0.4691 | 0.0338 | 0.0085 | 0.0548 |
| | SO | 1.3671E-01 | 0.4596 | 0.0345 | -0.0045 | 0.0399 |
| | NO | 5.6687E-01 | 0.4495 | 0.0346 | -0.0113 | 0.0204 |
| | IO | 2.5509E-01 | 0.4624 | 0.0317 | -0.0086 | 0.0337 |
| | TO | 1.4770E-01 | 0.4354 | 0.0277 | -0.0045 | 0.0349 |

### Supplementary Information Bibliography

1. Rupesh Agrawal, Preeti Gupta, Kara-Anne Tan, Chui Ming Gemmy Cheung, Tien-Yin Wong, and Ching-Yu Cheng. Choroidal vascularity index as a measure of vascular status of the choroid: measurements in healthy eyes from a population-based study. *Scientific reports*, 6(1):21090, 2016.
2. Bjorn Kaijun Betzler, Jianbin Ding, Xin Wei, Jia Min Lee, Dilraj S Grewal, Sharon Fekrat, Srinivas R Sadda, Marco A Zarbin, Aniruddha Agarwal, Vishali Gupta, et al. Choroidal vascularity index: a step towards software as a medical device. *British Journal of Ophthalmology*, 106(2):149–155, 2022.
3. Leslie G Portney. *Foundations of clinical research: applications to evidence-based practice*. FA Davis, 2020.
4. Xin Wei, Shozo Sonoda, Chitaranjan Mishra, Neha Khandelwal, Ramasamy Kim, Taiji Sakamoto, and Rupesh Agrawal. Comparison of choroidal vascularity markers on optical coherence tomography using two-image binarization techniques. *Investigative Ophthalmology & Visual Science*, 59(3):1206–1211, 2018.
